# Cilta-cel salvages ide-cel failure in relapsed multiple myeloma by driving distinct immune responses

**DOI:** 10.1101/2025.07.10.25331322

**Authors:** Tony Pan, Erting Tang, Yifei Hu, Nicholas Asby, Mckenzie Schubat, Thomas Althaus, Peter A. Riedell, Benjamin Derman, Jun Huang

## Abstract

Ide-cel and cilta-cel are the two FDA-approved anti-BCMA CAR T cell therapies for the treatment of relapsed/refractory multiple myeloma. Here, we studied a patient who was initially treated with ide-cel with progressive disease and subsequently treated with cilta-cel with a complete response. To elucidate the underlying mechanisms underpinning the distinct clinical outcomes, we conducted multimodal, cross-tissue, and longitudinal single-cell analyses. This enabled us to directly compare the specific cellular and molecular factors distinguishing these two CAR therapies including their cell phenotypes, post-infusion kinetics, and endogenous immune landscapes. We found that the ide-cel infusion product was dominated by CD4^+^ CAR T cells, upregulated a terminal effector phenotype, and exhibited elevated activation signatures. Post-infusion, ide-cel CAR T cells failed to proliferate, sustain cytotoxicity, or migrate into the bone marrow, resulting in persistent myeloma cells and dysregulated monocytes and natural killer cells. In contrast, the cilta-cel infusion product exhibited a balanced ratio of CD4^+^ and CD8^+^ CAR T cells, upregulated a resident memory-like signature, and displayed signatures of IL-1 and IL-2 family cytokine signaling. Post-infusion, cilta-cel CAR T cells retained their resident memory-like profile, were durably retained in the peripheral blood, and successfully infiltrated the bone marrow, leading to effective tumor clearance and reestablishment of immune homeostasis. Our results present important clinical evidence that cilta-cel can serve as an effective salvage treatment following ide-cel failure. By providing a direct patient-matched comparison between two CAR therapies, our study uncovers important insights into both CAR T-cell intrinsic properties and immune environmental factors that contribute to effective BCMA CAR T-cell treatment.

## INTRODUCTION

Multiple myeloma, arising from malignant plasma cells in the bone marrow, is the second-leading hematological malignancy and remains largely incurable despite recent therapeutic developments^1^. While modern myeloma treatment regimens have yielded improved response rates and progression-free survival durations^2–4^, most patients eventually relapse or become refractory to treatment.

Chimeric antigen receptor (CAR) T-cell therapy has emerged as a promising option for the treatment of relapsed/refractory multiple myeloma (RRMM). The two FDA-approved CAR T cell products, idecabtagene vicleucel (ide-cel) and ciltacabtagene autoleucel (cilta-cel), have substantially improved clinical treatment efficacy compared to standard-of-care in heavily pretreated RRMM patients^5,6^. Both CARs possess CD8α transmembrane and 4-1BB/CD3ζ intracellular domains, but they differ in their extracellular binding domains targeting B-cell maturation antigen (BCMA) expressed by myeloma cells. Ide-cel has an extracellular domain consisting of a single anti-BCMA scFv, whereas cilta-cel has an extracellular domain comprising two anti-BCMA nanobodies^7,8^. Prior studies comparing the clinical efficacy of cilta-cel versus ide-cel suggest that cilta-cel is associated with better treatment responses and longer progression-free survival^9,10^. However, ide-cel has a shorter median turnaround time between apheresis and infusion (47 days) compared to cilta-cel (68 days)^11^. Clinical guidelines for selecting between these two BCMA CAR therapies remain underdeveloped, partly because of the lack of data directly comparing their biological properties^12^. Additionally, the efficacy and benefits of using one BCMA CAR as salvage therapy following the failure of another remain largely unexplored^13^.

To address this unmet clinical need, we conducted comprehensive analyses on a unique case involving a patient with RRMM who was initially treated with ide-cel with progressive disease and subsequently treated with cilta-cel achieving a complete response. To understand the underlying mechanisms behind these distinct clinical outcomes, we performed multimodal (RNA, surface protein, TCR), cross-tissue (peripheral blood and bone marrow), and longitudinal (infusion product and day 9-28 post-infusion) single-cell analyses. Our analyses capture all phases of the CAR T-cell therapy cycle, beginning with initial infusion products, followed by CAR T-cell *in vivo* expansion, bone marrow infiltration, tumor cell eradication, and culminating in the reestablishment of immune homeostasis. These findings not only demonstrate that cilta-cel can effectively salvage ide-cel failure but also offer valuable insights to enhance CAR T cell functionality and persistence.

## RESULTS

### Clinical background and study design

The patient is a man in his late 50s with IgG-lambda myeloma and high-risk cytogenetics (Supplemental Table 1). At diagnosis, he received triplet therapy with bortezomib, lenalidomide, and dexamethasone, followed by tandem autologous stem cell transplantation and lenalidomide maintenance therapy. He achieved a best response of minimal residual disease (MRD) negative complete response (CR) before eventual relapse. He then received daratumumab, carfilzomib, and dexamethasone but relapsed less than a year later (**Fig**. 1a, Table 2). He subsequently received ide-cel CAR T cell therapy with fludarabine and cyclophosphamide lymphodepletion; he experienced grade 2 cytokine release syndrome (CRS) and received one dose of tocilizumab. He had progressive disease (PD) following ide-cel with new hypermetabolic tumor activity in his bilateral humeri (**Figs**. 1a-b). Following holding therapy with pomalidomide and dexamethasone, he was collected for and received salvage cilta-cel CAR T cell therapy with bendamustine lymphodepletion (due to fludarabine shortage); he had grade 1 CRS and immune-effector cell-associated hemophagocytic lymphohistiocytosis-like syndrome for which he received tocilizumab and dexamethasone (Supplemental Table 2). He achieved an MRD negative (10^-6^, next-generation sequencing) CR, with significant reduction of hypermetabolic lesions in his bilateral humeri and no evidence of new lesions (**Figs. 1a-b**) in addition to normalization of circulating IgG lambda (**Fig. 1c**).

**Fig. 1.**
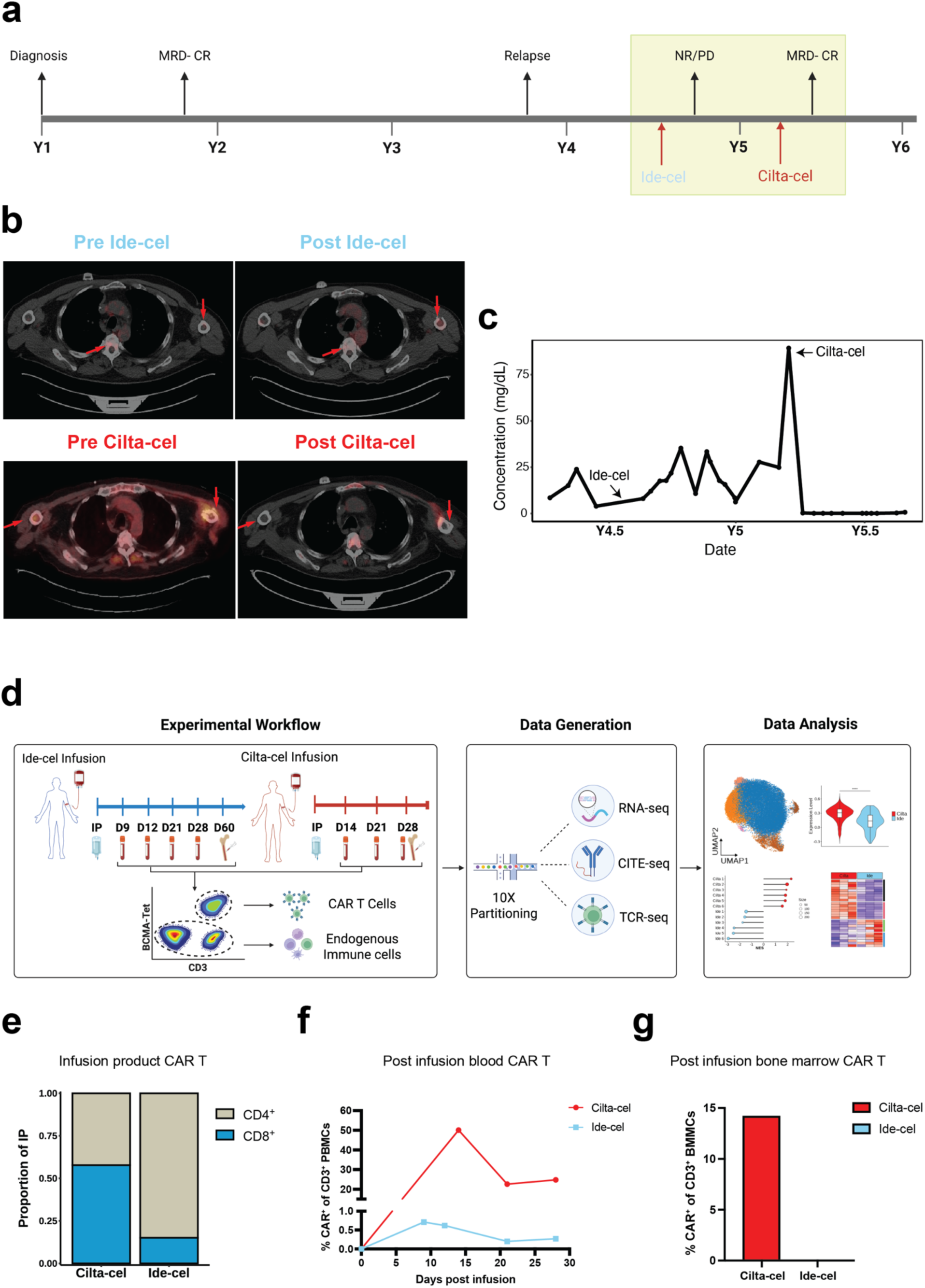
Cilta-cel and ide-cel elicit divergent clinical responses and display distinct post-infusion kinetics and infusion product compositions. **(a)** Timeline of the patient’s RRMM history and CAR T cell treatment. Highlighted region indicates the timeline of samples used in this study. **(b)** PET/CT images indicating hypermetabolic activity of the patient’s bilateral humeri before and after each CAR T cell treatment. Red arrows indicate regions where lesions were significantly altered following treatment. **(c)** Line plot denoting the patient’s peripheral blood concentration of IgG lambda light chain over time before and after each CAR T cell infusion. **(d)** Schematic illustrating the sample collection, sorting strategy, sequencing data generation, and data analysis workflow. **(e)** Bar plot indicating the phenotype proportion (CD4^+^ or CD8^+^) of each CAR T cell infusion product by sequencing analysis. **(f)** Line plots depicting the change in peripheral abundance of each CAR across each timepoint by flow cytometry. **(g)** Bar plot indicating the abundance of each CAR in the bone marrow by flow cytometry. Abbreviations: cilta, cilta-cel; ide, ide-cel; PET/CT, positron emission tomography/computed tomography; IgG, immunoglobulin G.

Given the distinct treatment responses observed in the same patient, we hypothesize that the markedly different outcomes between ide-cel and cilta-cel therapies can be attributed to variations in the CAR-T cell infusion products, post-infusion CAR-T cell differentiation, and/or the endogenous immune landscapes. To understand the underlying root causes and associated molecular mechanisms of the two distinct clinical outcomes, we performed longitudinal single-cell multimodal analyses (paired RNA-seq/CITE-seq/TCR-seq) of the patient’s CAR-T cell infusion products, post-infusion peripheral blood mononuclear cells (PBMCs), and post-infusion bone marrow biopsies. These analyses directly reveal the CAR-T cell phenotypic heterogeneity, differentiation trajectories, and clonal dynamics associated with each CAR-T cell therapy at the transcriptomic level (**Fig.** 1d). Post-infusion CAR T cells were sorted using BCMA-tetramers^14^ (**Fig.** S1). All CAR-negative immune cells were sequenced to interrogate the endogenous immune landscapes (**Fig**. S1).

We first found that both the composition and post-infusion abundance of CAR T cells varied substantially between the two therapies. Consistent with prior findings^15,16^, the ide-cel infusion product was CD4^+^-dominant (15% CD8^+^ and 85% CD4^+^) while the cilta-cel infusion product had a more balanced ratio of CD8^+^ and CD4^+^ CAR T cells (58% CD8^+^ and 42% CD4^+^) (**Fig.** 1e). Ide-cel and cilta-cel peak expansion occurred at D9 and D14 respectively, which is in line with previous studies^5,6,11^ (**Fig.** 1f). Ide-cel was minimally detected at both peak and post-peak (D12, D21, D28) timepoints (**Figs.** 1f), despite a consistent presence of T cells (**Fig.** S2). In contrast, cilta-cel was readily detected and persisted throughout day 21 and 28 in the peripheral blood (**Figs.** 1f). Furthermore, we investigated CAR T-cell infiltration in the bone marrow. Cilta-cel was readily detected whereas ide-cel was undetectable with a limited number of overall T cells observed (**Figs.** 1g, S3a-b), suggesting that cilta-cel, but not ide-cel, effectively infiltrated into the bone marrow for tumor cell killing.

### Cilta-cel and ide-cel infusion products exhibit distinct phenotypic and cytokine expression profiles

Prior research has indicated that intrinsic factors within CAR T-cell infusion products play a crucial role in determining treatment efficacy^17–19^. To examine whether variations in the infusion products underlie the observed differences in treatment outcomes, we conducted a detailed comparison of the cellular profiles of these two infusion products.

We first focused on CD8^+^ CAR T cells. Uniform Approximation and Projection (UMAP) dimensional reduction and clustering identified three CAR T-cell populations: proliferating effector memory (Prolif_EM, *MKI67*^hi^*SELL*^+^*GNLY*^-^), proliferating terminal effector (Prolif_TE, *MKI67*^hi^*SELL*^lo^*GNLY*^+^), and terminal effector (TE, *MKI67*^lo^*SELL*^-^*GNLY*^+^) (**Figs.** 2a-b). Proliferating effector memory cells expressed higher levels of memory-associated genes (*SELL, LEF1*) and proteins (CD127, CD62L, CD45RA) while proliferating terminal effector and terminal effector cells expressed higher levels of effector-associated genes (*GZMA, TNF, PRF1*) and proteins (KLRG1, CD45RO) (**Fig.** 2b). Importantly, cilta-cel and ide-cel CD8^+^ CAR T-cells had distinct compositions and generally clustered separately. Cilta-cel predominantly consisted of proliferating effector memory cells, while ide-cel primarily consisted of proliferating terminal effector and terminal effector cells (**Figs.** 2a bottom, S4a). Consistent with our phenotypic findings, cilta-cel CAR T cells upregulated memory-associated genes (including *SELL*, *LEF1*, and *FOXO1*) and proteins (**Figs**. 2c, S4b). Ide-cel CD8^+^ CAR T cells, however, upregulated inhibitory receptors (including *HAVCR2*, *LAG3*, *ENTPD1*) (**Figs.** 2c, S4c), in addition to effector-associated genes and proteins (including *GNLY*, *NKG7*, *GZMA*, *PRF1)* (**Figs**. S4d-e). We also confirmed that differences in memory-associated and inhibitory receptor gene expression were consistent across all three CD8^+^ populations (**Figs.** S4f-h), indicating globally distinct profiles.

**Fig. 2.**
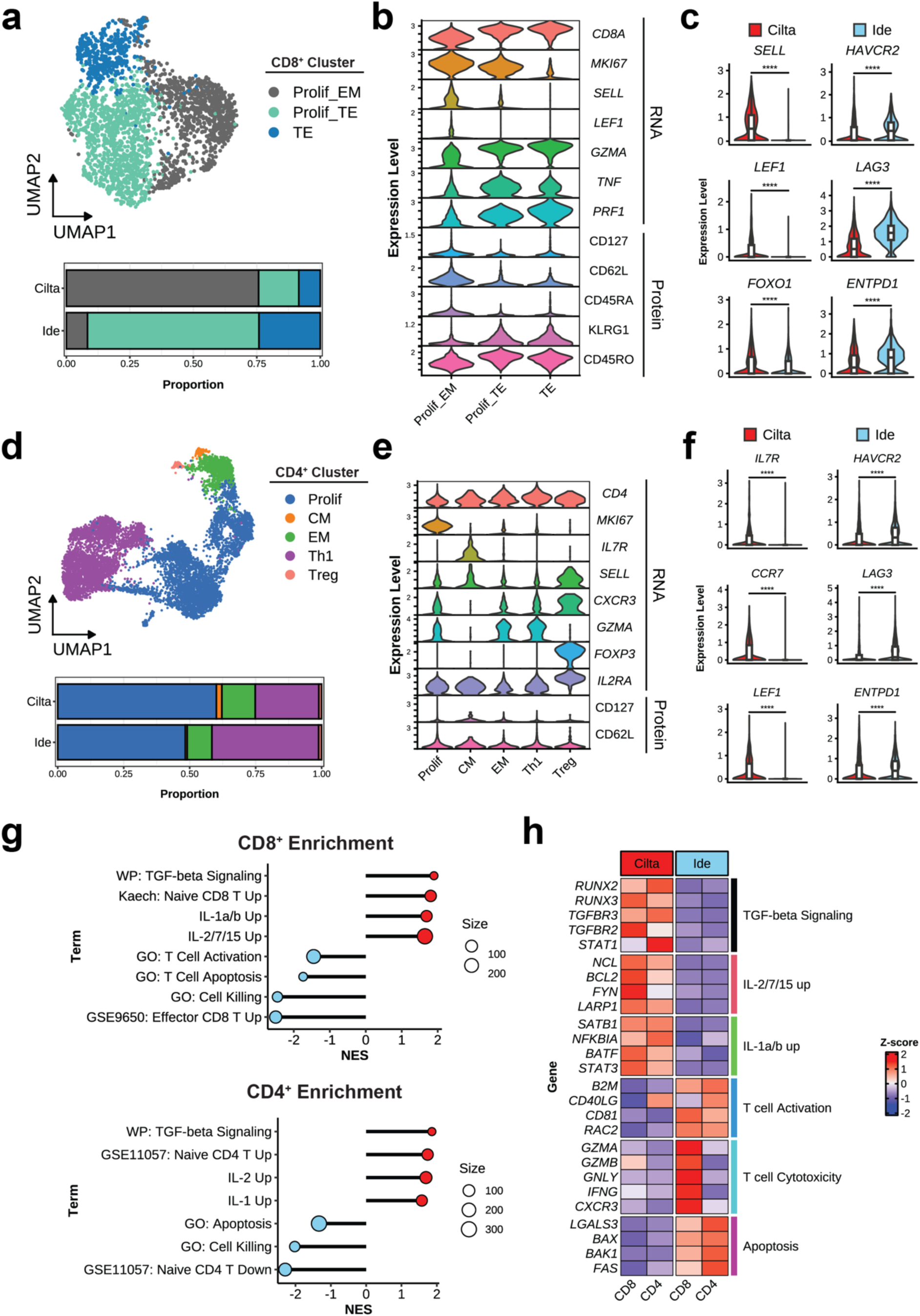
Phenotypic and transcriptomic signatures distinguish cilta-cel and ide-cel CAR T cell infusion products. (**a**) Top: UMAP plot of CD8^+^ CAR T cells colored by phenotypic cluster. Bottom: bar plot depicting the proportions of each subset for each therapy. (**b**) Violin plots depicting normalized expression levels of key genes and proteins used to annotate CAR T-cell phenotypes. (**c**) Violin plots contrasting memory-associated (left) and inhibitory receptor (right) gene expression of each treatment. Expression levels were compared by Wilcoxon rank-sum test with Bonferroni adjustment, whereby **** indicates p<.0001. (**d**) as in (**a**), but for CD4^+^ CAR T cells. (**e**) as in (**b**), but for CD4^+^ CAR T cells. (f) as in (c), but for CD4^+^ CAR T cells. (**g**) Dot plots depicting enriched pathways on DEGs upregulated by each treatment for CD8^+^ (top) and CD4^+^ (bottom) CAR T cells. (**h**) Heatmap depicting scaled expression of selected DEGs and their associated pathways, grouped by phenotype and split by treatment. Abbreviations: Prolif_EM, proliferating effector memory; Prolif_TE: proliferating terminal effector; TE: terminal effector; CM, central memory; EM, effector memory; Th1, T helper type 1; Treg, regulatory T cell, NES, normalized enrichment score; log_2_FC, log_2_fold-change.

We next asked if similar trends were present in infusion product CD4^+^ CAR T cells. Clustering identified 5 CAR T cell populations: proliferating (Prolif, *MKI67*^+^), central memory (CM, *IL7R*^hi^*SELL*^hi^), effector memory (EM, *IL7R*^-^*CXCR3*^lo^), T-helper type 1 (Th1, *IL7R*^-^*CXCR3*^hi^), and T regulatory (Treg, *FOXP3*^+^*IL2RA*^hi^) T cells (**Fig.** 2d-e). Although we did not observe as clear of a phenotypic shift (**Fig.** S5a), we similarly found a cilta-cel specific upregulation of memory-associated genes *(IL7R, CCR7*, *LEF1*) and proteins (**Fig.** 2f and S5b) compared to an ide-cel specific upregulation of inhibitory receptor and effector genes and proteins (**Figs.** S5c-e). While cilta-cel consistently upregulated memory-associated genes across the three largest clusters, ide-cel upregulation of inhibitory receptors was specific to the proliferating cluster, suggesting an elevated activation profile (**Fig.** S5f-h).

To further understand the molecular factors differentiating the infusion products, we performed differential gene expression (DEG) analysis and gene set enrichment analysis (GSEA). Across both CD8^+^ and CD4^+^ CAR T cells, cilta-cel upregulated naïve T cell signatures and distinct cytokine signaling pathways; TGF-β (*RUNX2*, *RUNX3*, *TGFBR1*, *TGFBR2*), which can induce a tissue resident memory T cell (TRM)-like phenotype^20,21^, IL-2 family (consisting of IL-2, IL-7, and IL-15) (*NCL*, *BCL2*, *FYN*, *LARP1*), which promote CAR T cell proliferation, effector function, and persistence^22,23,24^, and IL-1 family (consisting of IL-1α and IL-1β) (*SATB1*, *NFKBIA*, *BATF*, *STAT3)*, which enhances T cell antitumor immunity^25,26^ (**Figs**. 2g-h). On the other hand, ide-cel CAR T cell pathways were enriched in T cell activation (*B2M*, *CD40LG*, *CD81*) and killing (*GZMA, GZMB*, *GNLY*), but also apoptosis (*BAX, BAK1*) (**Figs**. 2g-h).

We next constructed gene regulatory networks and identified treatment-specific regulons for both CD8^+^ (**Figs**. S6a-b) and CD4^+^ CAR T cells (**Figs**. S6c-d). Consistently, top cilta-cel regulons were enriched in memory-associated regulons including *MYB, LEF1*, and *STAT1*^27–29^ (**Figs**. S6a-d). High-confidence target genes present in these regulons included *CCR7, NELL2, ETS1*, and *IRF4*, which drive T cell stemness, and *FYN*, which promotes T-cell signaling (**Fig.** S7a-b). Top ide-cel regulons were enriched in inflammatory and terminal differentiation-associated regulons including *TBX21*, *BHLHE40*, *RORC*, and *MAF* (**Figs**. S6a-d). High-confidence target genes present in these regulons included T-cell effector (including *KLRG1*, *GZMA*, *CCL5*, *CXCR3*, and *TNFAIP3*) and activation (including *PDCD1*, *TNFRSF1B*, and *BCAT1*) (**Fig.** S7a-b). Together, this patient’s contrasting infusion product signatures reinforce previous studies describing factors distinguishing favorable versus unfavorable responses to CD-19 CAR T cell therapy^17,30–34^.

### Cilta-cel infusion product CAR T cells exhibit a T resident memory-like profile

Upregulation of *RUNX3*, a key gene associated with the T resident memory (TRM) lineage^35^, for both CD8^+^ and CD4^+^ cilta-cel CAR T cells (**Fig.** 2h) led us to hypothesize that the cilta-cel infusion product was conditioned toward a TRM-like transcriptional profile. Notably, all the top genes present in the *RUNX3* regulon were upregulated by cilta-cel CAR T cells (**Fig.** 3a, left), which included TGF-β-associated genes as well as regulators of T-cell development and signaling (including *FYN*, *ETS1*, *ZBTB17,* and *STAT3*). In contrast, the majority of the top genes present (including *ANXA2*, *AQP3*, and *CDKN2A*) in the *KLF2* regulon (which opposes TRM formation^36^) were upregulated by ide-cel CAR T cells (**Fig.** 3a, right). Expression of the *RUNX3* and *KLF2* regulons were respectively highly specific to cilta-cel and ide-cel for both CD8^+^ (**Fig.** 3b, left) and CD4^+^ (**Fig.** 3b, right) CAR T cells, suggesting strong treatment-specific activation of the two transcription factors (TFs).

**Fig. 3.**
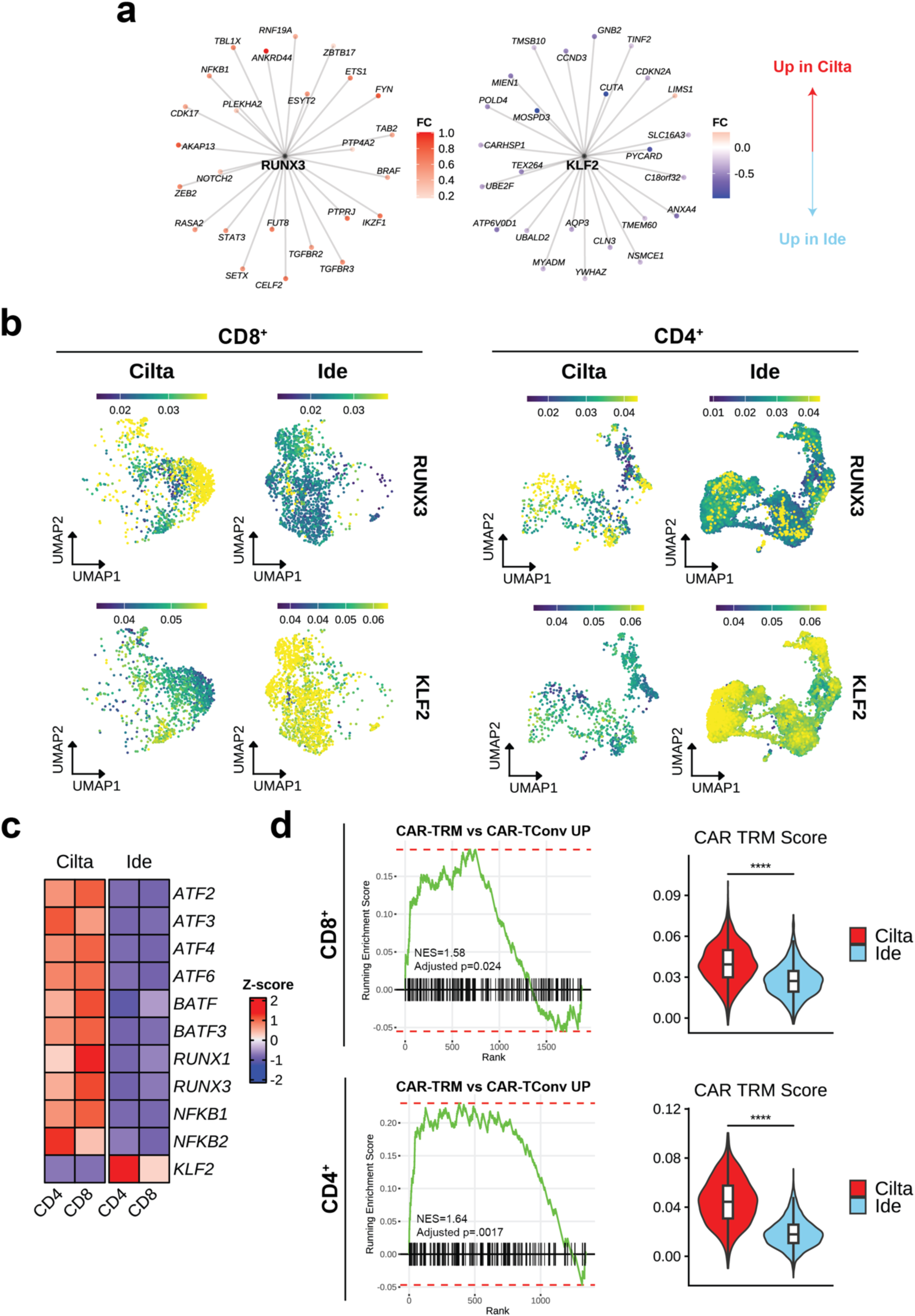
Cilta-cel infusion product CAR T cells display a TRM-like transcriptional signature. (**a**) Network plots of the *RUNX3* and *KLF2* regulon target genes. In each regulatory network, only the top weighted genes are depicted. Genes are colored according to overall log_2_fold-change between expression in cilta-cel (red) versus ide-cel (blue). (**b**) Split expression plots of the *RUNX3* (top) and *KLF2* (bottom) regulon for CD8^+^ (left) and CD4^+^ (right) CAR T cells. (**c**) Heatmap depicting scaled expression of differentially accessible CAR-TRM transcription factors from Jung et al., grouped by phenotype and split by treatment. (**d**) Left: enrichment plot of the CAR-TRM signature from Jung et al. on DEGs between cilta-cel and ide-cel in CD8^+^ (top) and CD4^+^ (bottom) CAR T cells. Right: Violin plots contrasting expression of the CAR-TRM signature from Jung et al. in CD8^+^ (top) and CD4^+^ (bottom) CAR T cells. Expression levels were compared by Wilcoxon rank-sum test with Bonferroni adjustment, whereby **** indicates p<.0001. Abbreviations: CAR-TRM, resident memory CAR T cell; CAR TConv, conventional CAR T cell.

We next cross-referenced our data with existing literature on TRM transcriptomic profiles. A recent study by Jung et al. reported that CAR T cells treated with TGF-β upregulate a TRM-like (CAR-TRM) transcriptional and epigenetic profile and demonstrate enhanced efficacy in treating blood and solid cancers compared to conventional CAR T cells^37^. We first confirmed that cilta-cel CAR T cells differentially upregulated CAR-TRM accessible TFs from Jung et al. and downregulated *KLF2* (**Fig.** 3c). GSEA on the CAR-TRM gene signature demonstrated significant positive enrichment by cilta-cel CD8^+^ (p = 0.024) and CD4^+^ CAR T cells (p < .001) (**Fig.** 3d, left). Furthermore, cilta-cel CAR T cells overall expressed an elevated CAR-TRM gene signature compared to ide-cel CAR T cells (**Fig.** 3d, right). In summary, the elevated TRM-like profile may have facilitated cilta-cel CAR T-cell persistence and bone marrow infiltration post-infusion.

### CAR-TRM profiles persist in post-infusion cilta-cel CD8^+^ CAR T cells

We next investigated dynamics of post-infusion CAR T cells by tracking persisting CAR T-cell clones. We first assessed the infusion product clonal repertoire and found that the size distribution was similar between treatments (**Fig.** S8a). Notably, nearly all large clones (clones that occupy greater than 1% of the repertoire) for both therapies were CD8-positive (**Fig.** 4a). However, while large cilta-cel infusion product clones were highly expanded (8997 recovered TCRs at D14) and persistent in every post-infusion sample (average of 7015 recovered TCRs from D21-D28) (**Fig.** 4b, top), large ide-cel infusion product clones had minimal expansion (92 recovered TCRs at D9) or persistence (16 recovered TCRs at D28) (**Fig.** 4b, bottom). Clonal repertoires between cilta-cel and ide-cel did not overlap (**Fig**. S8a).

**Fig. 4.**
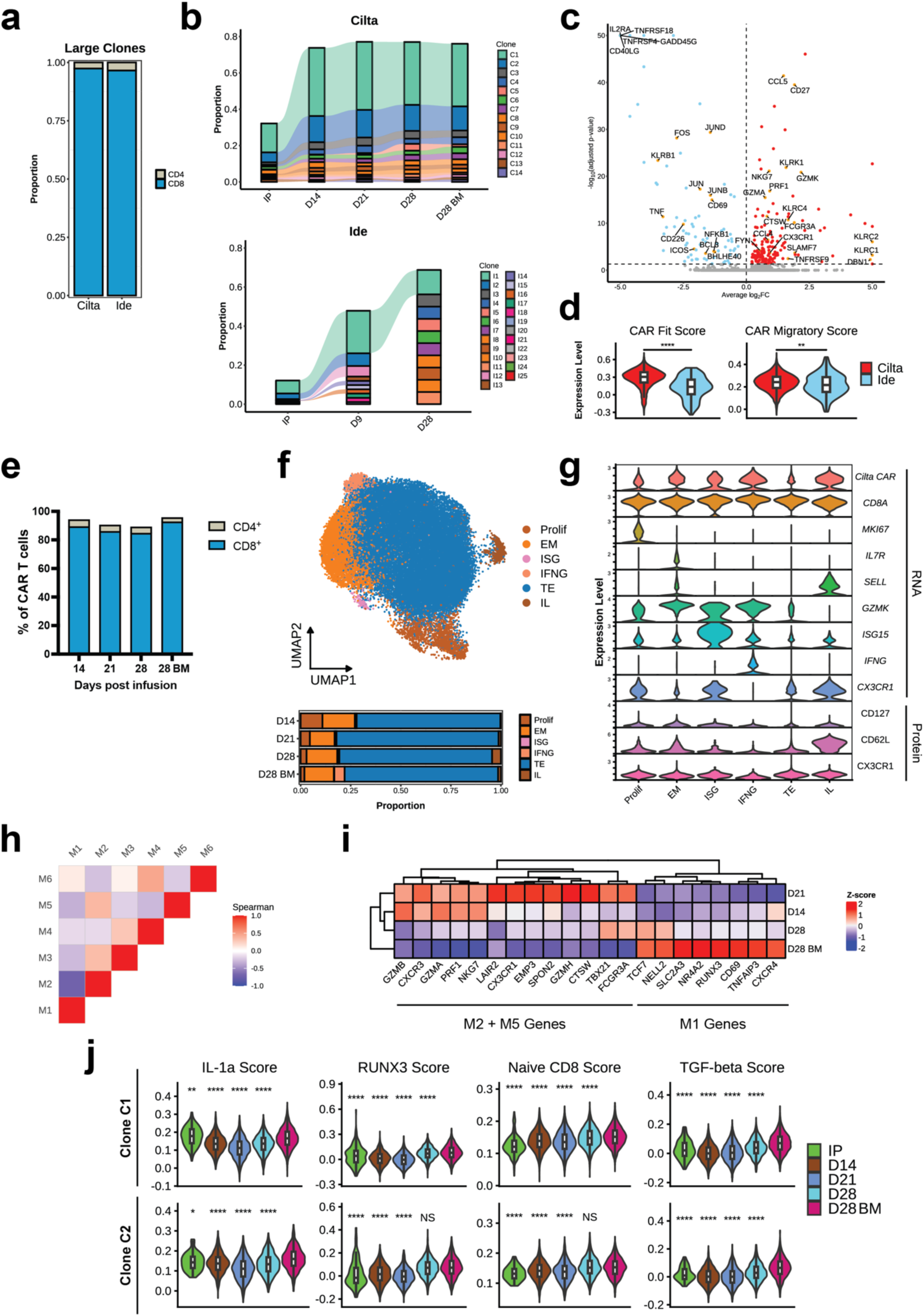
Post-infusion cilta-cel CAR T cells retain cytokine signatures and display enhanced cytotoxicity and persistence. (**a**) Distribution of large clones across CD8^+^ and CD4^+^ phenotypes. Large clones are defined as clonotypes that occupied greater than 1% of a sample’s TCR repertoire. (**b**) Alluvial plots depicting the abundance of the top 10 largest cilta-cel (top) and ide-cel (bottom) CAR T-cell clones at each timepoint. (**c**) Volcano plots of DEGs expressed by cilta-cel (right) and ide-cel (left) CAR T cells at peak expansion. Significant genes are colored according to log_2_fold-change between expression in cilta-cel (red) and ide-cel (blue). Selected genes are highlighted in orange. (**d**) Violin plots contrasting expression of CAR fitness and migration signatures from Rezvan et al. Expression levels were compared by Wilcoxon rank-sum test with Bonferroni adjustment, whereby **** indicates p<.0001 and ** indicates p <.01. (**e**) Bar plots depicting the phenotypic proportion (CD4^+^ or CD8^+^) of CAR T cells at each post-infusion timepoint as determined by flow cytometry. (**f**) Top: UMAP plot of CAR T cells colored by cluster. Bottom: bar plot depicting the proportion of each cluster at each timepoint. (**g**) Violin plots depicting normalized expression levels of genes used to annotate each cluster. (**h**) Heatmap depicting spearman correlation between gene modules identified in the TE cluster. (**i**) Heatmap depicting scaled expression of selected M1, M2, and M5 genes across timepoints. Timepoints are clustered based on Euclidean distance. (**j**) Violin plots depicting the expression of the IL-1a, *RUNX3* regulon, naïve CD8^+^, and TGF-β gene signature scores at each timepoint for clone C1 (top) and C2 (bottom). All significance comparisons are made relative to the D28 BM timepoint using the using the Wilcoxon rank-sum test with Bonferroni adjustment, whereby **** indicates p<.0001, * indicates p<.05, and ns indicates not significant.

We contrasted the gene expression profiles at peak expansion to interrogate the difference between peak-expanded cilta-cel and ide-cel CAR T cells. We found that both CAR T cells significantly upregulated T-cell activation markers; ide-cel expressed higher *TNFRSF4*, *TNFRSF18*, *CD40LG*, *ICOS*, and *CD69* while cilta-cel expressed higher *CD27*, *TNFRSF9*, *SLAMF7*, and *FYN* (**Fig.** 4c). However, many cytotoxic genes initially upregulated by ide-cel in the infusion product became downregulated at peak expansion, including *GZMA*, *PRF1*, *NKG7*, and *CCL5* (**Fig.** 4c). Cilta-cel CAR T cells additionally expressed markers of mature effector development, including *FCGR3A, CX3CR1, KLRC1,* and *KLRC2* (**Fig.** 4c). We confirmed that peak cilta-cel CAR T cells overall upregulated the serial killing (CAR fitness) and migratory gene signature of multifunctional CD8^+^ CAR T cells associated with superior clinical efficacy^38^ (**Fig.** 4d). This distinct contrast to signatures in the infusion product demonstrates a distinct pattern of effector function between the two therapies: ide-cel cytotoxicity appears to have peaked in the infusion product and became diminished following infusion, whereas cilta-cel cytotoxicity was unleashed to coincide with peak expansion.

We next investigated the phenotypic and transcriptional signatures of persistent cilta-cel CAR T cells. Because post-infusion CAR T cells were primarily CD8^+^ (**Fig.** 4f), we altogether clustered all CD8^+^ CAR T cells from both the peripheral blood and bone marrow from early (D14 and D21) and late (D28 and D28 BM) timepoints. We found 6 subsets: proliferating (*MKI67^+^)*, EM (*IL7R*^+^*GZMK^hi^*), interferon-stimulated (ISG, *ISG15^+^*), interferon-gamma-high (IFNG, *IFNG^+^)*, TE (*CX3CR1^hi^IL7R^-^)* and innate-like (IL, *TYROBP^+^*) (**Figs.** 4f-g). In contrast to previous studies of *in vivo* anti-CD19/CD28 CAR T-cell differentiation^39^, cluster composition was not biased by timepoint or clone size (**Figs**. 4f, S8b-c). We performed weighted gene co-expression network analysis (WCGNA) on the dominant TE subset to decompose temporal CAR T-cell gene expression into correlated gene programs. Among the 6 modules (M) discovered, we found that M1 (consisting of TRM-associated genes such as *NR4A2, CXCR4*, *CD69*, *RUNX3,* **Fig.** S9) was anticorrelated with M2 and M5 (**Fig.** 4h), which consisted of cytotoxic genes such as *PRF1*, *GZMB*, *GZMA*, *CX3CR1, FCGR3A* (**Figs.** S10-11). We found that that M1 genes were most highly expressed by D28 BM CAR T cells, which confirms their TRM-like nature (**Fig.** 4i).

The D28-specific upregulation of TRM-associated genes led us to postulate that persisting late-timepoint cilta-cel CAR T cells retain their cytokine-induced TRM-like signatures induced in the infusion product. To confirm this, we tracked gene signature expression of the most repertoire-dominant clones (C1 and C2, **Fig**. 4b top), which respectively occupy 15.9% and 5.4% of the infusion product repertoire, at each timepoint (**Fig.** 4j). While IL-2 signature expression was more infusion-product specific **(Fig.** S12), the expression of IL1-a, *RUNX3* regulon, naïve CD8, and TGF-β signatures were durably retained throughout D28 for both C1 and C2 (**Fig.** 4j). Furthermore, expression of the C1 *RUNX3* regulon, naïve CD8, and TGF-β signatures for were the highest in bone marrow D28 CAR T cells (**Fig.** 4j). Altogether, these results reinforce that the superior ability of late-stage cilta-cel CAR T cells to persist and home to the bone marrow can be attributed to the maintenance of a TRM-like signature.

### Cilta-cel clears persistent tumor and reverses post-ide-cel inflammation in the bone marrow

The distinct abundance and TRM-like profile of cilta-cel CAR T cells in the bone marrow led us to hypothesize that the patient’s bone marrow microenvironment was differentially remodeled in response to each CAR T-cell therapy. Thus, we next sought to contrast the patient’s bone marrow immune landscape following each treatment. Out of the cell types observed, only the post-ide-cel bone marrow harbored plasma cells (**Figs**. 5a, S13a). We re-clustered all bone marrow B-lineage (B and plasma) cells and performed copy number variation analysis to determine the majority were neoplastic based on their aneuploidy probability (**Fig**. 5b). We further confirmed that all neoplastic B-lineage cells were solely present post-ide-cel and were eliminated post-cilta-cel (**Figs**. 5b, S13c), demonstrating effective cilta-cel mediated clearance of the myeloma tumor. A closer examination of marker genes revealed that the normal B-lineage cluster expressed *MS4A1* and *CD19* and were likely nonmalignant B cells (**Fig**. 5c). The tumor cluster expressed conventional plasma markers *SDC1* and *GPRC5D* in addition to *TNFRSF17* (**Fig**. 5c), which indicates the patient did not experience loss of BCMA antigen, which can drive CAR T cell failure^40^. However, the tumor did not express CD38 (**Fig**. 5c), which is typical for patients who have recently received anti CD38 monoclonal antibody therapy.

**Fig. 5.**
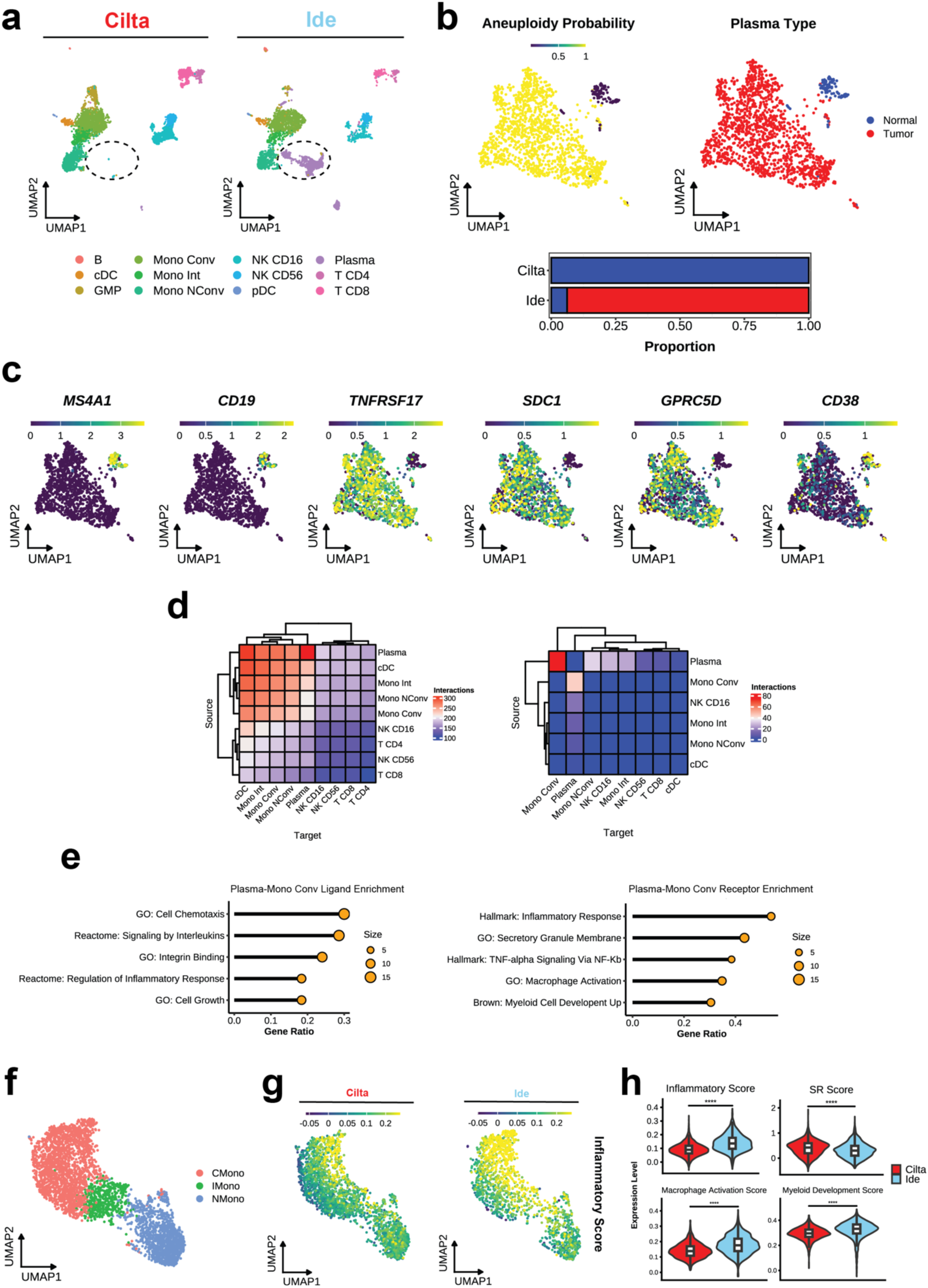
Cilta-cel and ide-cel infusions result in disparate endogenous immune landscapes in the bone marrow. (**a**) UMAP plot of the endogenous immune populations found in the bone marrow following CAR T cell infusion, colored by subset and split by treatment. Circled region indicates populations with treatment-specific enrichment. (**b**) UMAP plot of B-lineage cells in the bone marrow following CAR T cell infusion, colored by probability of tumor aneuploidy (left) and tumor status (right). Bottom: Bar plot depicting the proportion of tumor cells out of all B-lineage cells following each treatment. (**c**) Expression plot of key B-lineage genes in B-lineage cells. (**d**) Heatmaps demonstrating the number of putative interactions upregulated by all post-ide-cel subsets (left) and specifically by plasma cells (right). Rows depict source cells (expressing ligands), and columns depict target cells (expressing receptors). Rows and columns are clustered by Euclidean distance. (**e**) Dot plots depicting pathways enriched on plasma cell ligands that putatively interact with conventional monocyte receptors (left), and pathways enriched on conventional monocyte receptors that putatively interact with plasma cell ligands (right). (**f**) UMAP plot of post-infusion endogenous monocytes, colored by phenotype. (**g**) Expression plots of the Hallmark inflammatory gene signature in monocytes, split across treatments. (**h**) Expression plots of the inflammatory, SR monocyte, macrophage activation, and myeloid development gene signatures between treatments in monocytes. Abbreviations: CMono, conventional monocyte; IMono, intermediate monocyte; NMono, nonconventional monocyte; cDC, conventional dendritic cell; pDC, plasmacytoid dendritic cell; SR, standard risk.

To understand how post-ide-cel plasma cells putatively influenced the immune cellular communication network, we performed ligand receptor analysis between each cell subset and filtered for interactions that were specifically upregulated post-ide-cel. We found a high number of ligand receptor pairings between myeloid subsets and plasma cells (**Fig**. 5d, left); specifically, plasma cells most frequently upregulated ligands binding to receptors on conventional monocytes (**Fig**. 5d, right). We further investigated the immunomodulatory effects of plasma cells by applying gene set overrepresentation analysis to these interactions. Plasma cell ligands were enriched in pathways involving cellular development and communication (**Fig**. 5e, left) while monocyte receptors were enriched in pathways involving activation, development, and inflammatory signaling (**Fig. 5e**, right), suggesting that plasma cells stimulate monocytes toward a heightened inflammatory phenotype.

To more closely examine the differences in post-cilta-cel and post-ide-cel monocyte signatures, we separately clustered bone marrow monocytes and annotated subsets based upon *CD14* and *FCGR3A* expression (**Figs.** 5f, S14a-b). Consistent with previous studies associating elevated monocyte inflammation with poor MM outcomes^42,43^, we confirmed that post-ide-cel monocytes exhibited heightened inflammatory genes including *C3AR1*, *NAMP3*, *TLR2*, *IL4R*, and *IL1B* (**Figs.** 5g, S14c). In contrast, post-cilta-cel monocytes upregulated a standard-risk (SR) versus high-risk monocyte signature from Pilcher et al. (**Fig.** 5h, top), further confirming the polarization of post-ide-cel monocytes toward a poor prognostic transcriptional program. Post-ide-cel monocytes also exhibited elevated activation and maturation programs (**Fig.** 5h, bottom), which have been associated with elevated myeloma tumor burden^44,45^. To determine the genes associated with monocyte maturation, we inferred a gene-based trajectory of monocyte differentiation and found 3 trajectories (**Fig.** S14d). The T1 trajectory was initiated as the starting node as it included early-stage conventional monocyte genes such as *CD14*, *CCR2*, and *SELL* (**Fig.** S14d). T2 genes included nonclassical markers *CX3CR1*, *CXCL16*, and *FCGR3A* while T3 genes included *FCGR3B*, *VMO1*, and *SH2D1B* (**Fig.** S14d). We found that post-ide-cel monocytes expressed higher levels of T2 and T3 gene expression (**Fig.** S14e), confirming their polarization toward a more mature, nonclassical-like differentiation state. These distinct post-ide-cel signatures altogether indicate myeloma-induced monocyte dysfunction in the bone marrow.

To determine whether peripheral blood monocytes exhibited similar inflammatory signatures, we further interrogated monocytes in the peripheral blood (**Figs.** S15a-b). We found that one conventional monocyte subset (CMono3) and one intermediate monocyte subset (Imono2) were predominantly present post-ide-cel (**Figs.** S15c-d). Compared to other monocyte subsets, CMono3 and Imono2 highly upregulated inflammatory cytokines and chemokines (**Fig.** S15e). In agreement with bone marrow post-ide-cel monocyte signatures, we found a post-ide-cel-specific upregulation of inflammatory signaling and a downregulation of the SR gene set (**Fig.** S15f). GSEA analysis on CMono3 and IMono2 cluster-specific DEGs further revealed a shared theme of cytokine-driven inflammatory pathways (**Figs.** S15g-h). Together, these results point toward a systemically inflammatory post-ide-cel monocyte program.

### Cilta-cel restores dysfunctional post-ide-cell loss of NK-cell cytotoxicity

In addition to B-lineage cells and monocytes, we also identified a high abundance of NK cells in the bone marrow (**Figs**. 5, S13a). Clustering of the NK population revealed the presence of canonical CD56^hi^ and CD16^hi^ NK cells (**Figs.** 6a-b). CD16^hi^ NK cells expressed high levels of TNF signaling and cytotoxicity (**Fig.** 6c). We found that post-ide-cel NK cells were enriched in the CD56^hi^ subset and depleted in the mature CD16^hi^CX3CR1^hi^ cytotoxic subset compared to post-cilta-cel NK cells (**Fig.** 6a), a phenomenon which has previously been associated with poor myeloma prognosis^46^. Furthermore, post-ide-cel NK cells expressed lower levels of *FCGR3A* and higher levels of *NCAM1* (**Fig.** 6d). Post-ide-cel CD16^hi^ NK subsets also downregulated NK maturation and cytotoxicity genes including *SPON2*, *FGFBP2*, *PRF1*, and *GZMB* compared to post-cilta-cel NK cells (**Fig.** 6e). In the peripheral NK compartment (**Figs.** S16a-b), we again observed that post-ide-cel NK cells were consistently enriched in the CD56^hi^ subset and depleted in the CD16^hi^ subset compared to post-cilta-cel NK cells (**Figs.** S16c-d). Furthermore, we found that post-ide-cel NK cells expressed lower *FCGR3A* and the adhesion gene *CD226* in addition to downregulating the TNF and NK-mediated cytotoxicity pathways (**Fig.** S16e). We further explored differences in cytotoxic gene expression specifically in the CD16^hi^ subset and found similar downregulation by post-ide-cel NK cells compared to post-cilta-cel NK cells (**Fig.** S16f). Together, these results indicate that post-ide-cel NK cells experience dysregulation via both loss of the CD16^hi^ NK phenotype and CD16^hi^-mediated cytotoxicity.

**Figure 6:**
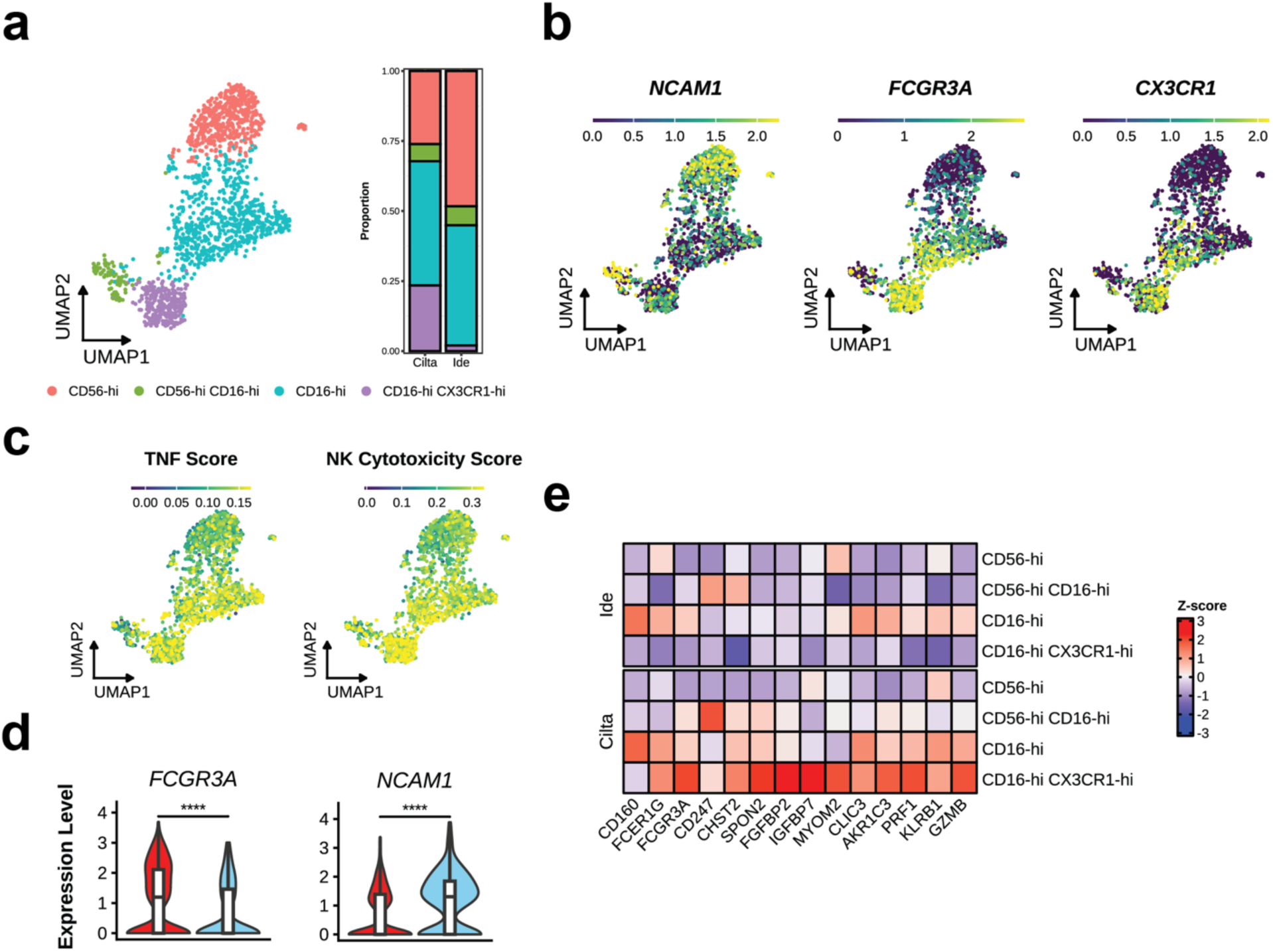
Phenotypic and transcriptomic characteristics of post-infusion NK cells in the bone marrow. (**a**) UMAP plot of NK cells, colored by phenotype (left); bar plot indicating the phenotype proportion for each treatment (right). (**b**) Expression plots of key genes used to annotate NK phenotypes. (**c**) Expression plots of TNF and NK cytotoxicity gene signatures. (**d**) Violin plots of *FCGR3A* and *NCAM1* for each treatment in NK cells. Expression levels were compared by Wilcoxon rank-sum test with Bonferroni adjustment, whereby **** indicates p<.0001. (**e**) Heatmap depicting scaled expression of selected cytotoxicity-associated DEGs in NK cells, grouped by subset and split by treatment. Abbreviations: TNF, tumor necrosis factor.

## DISCUSSION

Despite the significant clinical progress achieved by BCMA CAR T-cell therapies to treat RRMM, managing patients who fail CAR T-cell treatment remains a critical challenge in the clinic due to poor prognoses^47^. While various salvage therapy options including antibody-drug conjugates, T-cell-engaging therapies, and autologous stem cell transplants have been explored, mixed outcomes have been reported and the optimal sequencing or combination of therapies remains unclear^48^. Notably, the use of one commercial BCMA CAR T-cell therapy to salvage another has been minimally investigated. In this study, we performed longitudinal single-cell RNA-seq, CITE-seq, and TCR-seq on a unique clinical case of a patient with RRMM who received two successive FDA-approved anti-BCMA CAR T-cell therapies, ide-cel and cilta-cel. These two therapies yielded distinct outcomes of progressive disease and MRD-negative complete response in addition to divergent immune dynamics, summarized in Figure 7. Our findings suggest that cilta-cel can be a viable therapeutic option for patients who fail ide-cel, addressing an important gap in the current clinical understanding of BCMA CAR T-cell therapy.

**Fig. 7.**
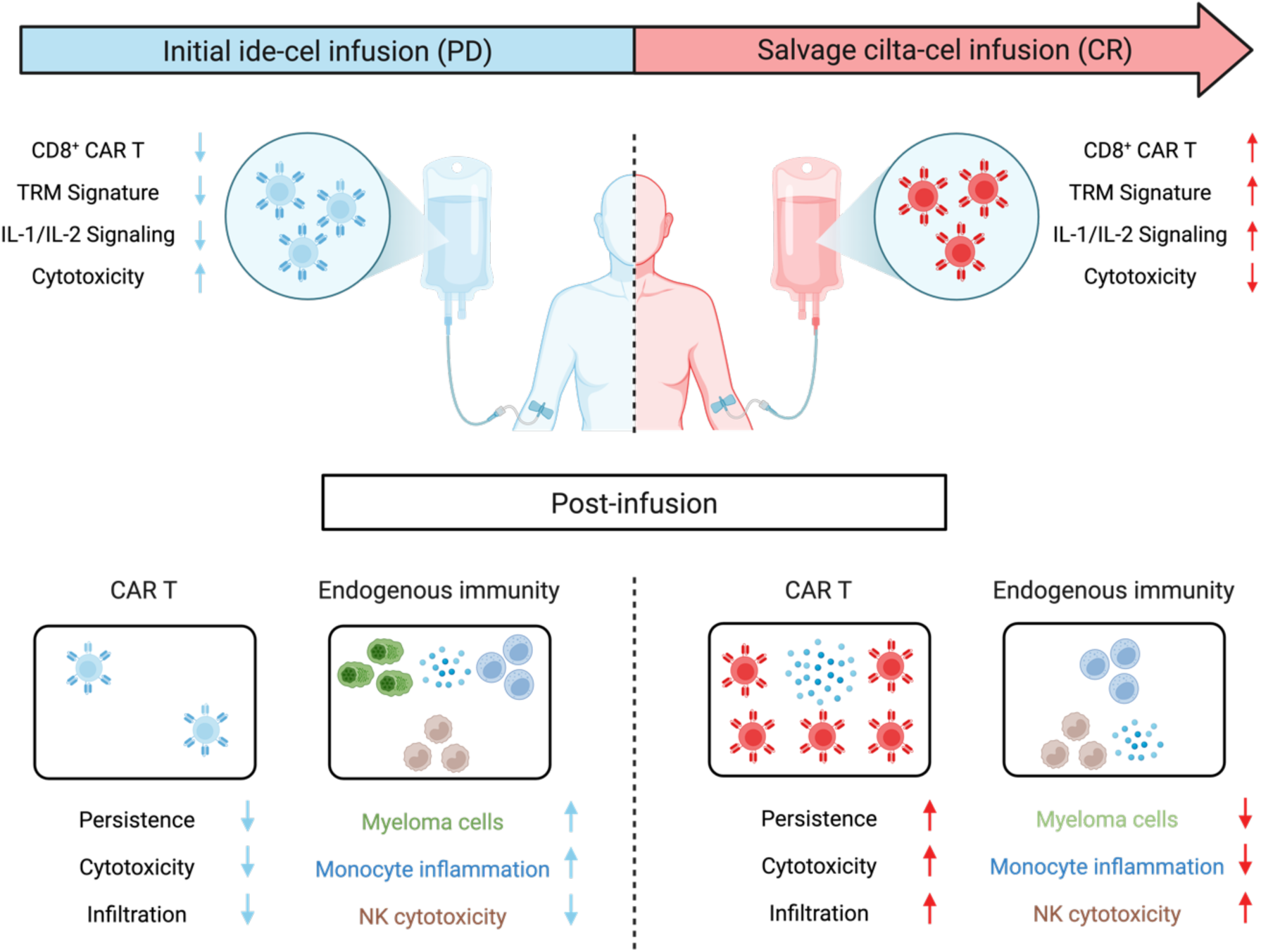
Summary of the infusion and post-infusion immune responses distinguishing cilta-cel and ide-cel. Elevated cytotoxicity and diminished CD8^+^ CAR T cells in the ide-cel infusion product leads to poor CAR T-cell persistence and cytotoxicity, resulting in persistent myeloma and dysregulated endogenous immunity. In contrast, an elevated TRM-like and IL-1/IL-2 cilta-cel infusion product signature contributes to durable CAR T-cell persistence and cytotoxicity, resulting in complete clearance of myeloma and homeostatic endogenous immunity.

While the patient’s superior response to cilta-cel is consistent with previous reports of superior clinical efficacy over ide-cel^5,6,49–51^, the underlying mechanisms remain unclear. We postulate that these differences stem from each therapy’s distinct infusion product phenotypic compositions and signatures. While the CD4^+^-dominant ide-cel infusion product was polarized toward a terminal effector-like phenotype with elevated expression of inhibitory receptors, the CD8^+^/CD4^+^ balanced cilta-cel infusion product was polarized toward a TRM-like phenotype that upregulated and sustained TGF-β and IL-1/IL-2 family cytokine signaling. These results are consistent with previous studies reporting that less activated, less differentiated infusion product phenotypes correlate with greater expansion, longer persistence, and overall more favorable therapeutic responses^17–19^, which is frequently mediated by IL-2, IL-7, and IL-15^17–19,52–54^. We also discovered that IL-1 and TGF-β signaling genes (including *ZEB2, TGFBR2, TGFBR3, RUNX3, NFKBIA*) overlapped with the TRM-determining *RUNX3* regulon, suggesting that IL-1 and TGF-β (which have both been shown to induce TRM differentiation and maintenance^21,37,55,56^) may have been complementarily utilized to drive a durable CAR TRM phenotype. Furthermore, the retention of the *RUNX3*, IL-1, and TGF-β signatures in D28 bone marrow CAR T cells suggests that the combination of IL-2 family and TRM-associated cytokine conditioning may be important for durable CAR T cell retention and efficacy in the treatment of RRMM.

In contrast, the simultaneous upregulation of effector and inhibitory markers and high proportion of CD4^+^ CAR T cells in the ide-cel infusion product are characteristic of elevated stimulation, which increases CD4/CD8 CAR T-cell ratios and exhaustion while decreasing expansion^57^. This short-lived effector cell phenotype, combined with increased apoptotic gene expression, likely explains the minimal post-infusion ide-cel expansion and persistence. As ide-cel non-responders have been shown to display inferior CAR T-cell expansion and persistence compared to responders^49,58^, the patient’s lack of ide-cel expansion and persistence past day 9 after infusion was likely a key determinant of response. Given that expression of inhibitory markers, increased activation, and more mature differentiation have been associated with poor efficacy due to loss of effector function, T-cell exhaustion, and poor expansion due to apoptosis^31,59,60^, our results are supported by prior studies and altogether highlight the link between BCMA CAR T-cell infusion product activation and treatment outcome.

The difference in tumor-clearing capability between ide-cel and cilta-cel may also be attributable to their antigen-binding domain design. Analysis of the bone marrow revealed the presence of *TNFRSF17*^+^*CD38*^-^ myeloma cells that were unable to be cleared by ide-cel. Notably, the lack of *TNFRSF17* expression or detected aneuploidy indicates that these myeloma cells were completely cleared by cilta-cel. Cilta-cel’s enhanced on-tumor targeting may be attributed to its dual VHH nanobody-based antigen-binding domains, which concurrently recognize two distinct BCMA epitopes. This dual-targeted design both increases binding affinity and decreases the risk of antigen escape. Conversely, ide-cel’s single-epitope scFv binding domain only engages one BCMA epitope, potentially limiting its capability to eliminate heterogenous tumor populations. These findings highlight the need to further investigate the optimal deployment of BCMA CAR T-cell therapies for specific subtypes of RRMM.

We also found that the persistent myeloma tumor elicited dysregulated post-ide-cel immune responses in both the peripheral blood and bone marrow. Myeloma cells contributed to elevated post-ide-cel monocyte inflammation, which has been associated with impaired CAR T-cell response as a result of excessive cytokine secretion^61,62^. Furthermore, the decrease of both the abundance and cytotoxic gene signature of post-ide-cel CD16^hi^ NK cells suggests that chronic myeloma cell activation may be inducing a dysfunctional NK profile lacking functional antitumor and immune surveillance. Importantly, we demonstrate that cilta-cel was able to overcome these immunological barriers to effectively clear myeloma cells and re-establish a homeostatic immune state.

Our study provides several important clinical implications. Firstly, our successful case of utilizing cilta-cel to salvage ide-cel failure points to a new frontier for BCMA CAR T-cell therapy. Clinical studies have reported that CR rates to a second CD19 CAR T-cell therapy in patients who failed an initial line of CD19 CAR T-cell therapy are typically low, ranging from only 20-30%^63,64^. Together with a lack of similar studies for BCMA CAR T-cell therapies and the high cost of administrating two separate CAR T-cell products, new clinical trials specifically investigating and optimizing BCMA CAR T-cell salvage therapy are critical for RRMM patient outcomes. Secondly, our results highlight the importance of BCMA CAR T-cell therapy selection, especially since each successive line of MM therapy decreases subsequent treatment efficacy^65^.

Despite the superior efficacy and response depth of cilta-cel compared to ide-cel in the treatment of RRMM, other clinical factors can complicate the choice between the two BCMA CAR T-cell therapies. For example, patients receiving cilta-cel typically experienced longer manufacturing times^66^, which may be problematic for patients with highly aggressive disease. Cilta-cel patients also experienced more severe side effects including hemophagocytic lymphohistiocytosis, enterocolitis^67^, and delayed motor and neurocognitive toxicities^66^, which can be fatal. Finally, our results suggest that cytokine-mediated promotion of a TRM-like phenotype is a potential strategy to induce superior CAR T-cell response, expansion, and persistence. This strategy may particularly benefit other hematological malignancies originating from the bone marrow such as leukemia. However, the clinical underperformance of CAR T-cell therapy (including cilta-cel) for RRMM patients with extramedullary disease^68,69^ suggests that additional site-specific barriers could be preventing sufficient infiltration, which warrants further investigation.

## METHODS

### Collection of patient biospecimens

Deidentified patient biospecimens were gathered in accordance with ethical guidelines under the institutional review board at the University of Chicago Medicine. Residual infusion product cells were collected from the patient’s infusion product bag and stored in freezing solution (Human serum albumin, 10% DMSO). Post-infusion PBMCs were collected from peripheral blood biospecimens by Ficoll-Plaque PLUS (Cytiva, 95021-205) and stored in freezing solution (PBS, 10% DMSO). Post-infusion bone marrow biopsies were collected from patient’s bone marrow aspirate and stored in freezing solution (PBS, 10% DMSO).

### Generation of BCMA-tetramers

Tetramers were constructed from AviTag-biotinylated human His-tagged BCMA/TNFRSF17 Protein (Acro Biosystems, BCA-H82E4) and streptavidin as previously described^14^. In brief, biotinylated BCMA was added to the tetrameric streptavidin at a 4:1 molar ratio for 30 minutes at 4°C in the dark. This mixture was diluted with PBS to convenient concentrations for staining. Alexa Fluor 647-labeled streptavidin (BioLegend, 405237) was used to prepare BCMA-tetramers for staining PBMCs and bone marrow biopsies.

### Single-cell RNA-seq/CITE-seq/TCR-seq

Cryopreserved biospecimens were thawed (RPMI, 10% FBS) and washed with cold FACS buffer (PBS, 2% BSA, 0.05% sodium azide). Fc receptors were blocked with Human TruStain FcX (BioLegend, 422301) at 1:50 dilution for 5 minutes at 4°C. Then, cells were incubated for 30 minutes at 4°C in the dark with a staining solution containing CITE-seq antibodies (described below), phenotyping antibodies (described below), and BCMA-tetramers (5 nM final concentration) for CAR T cell phenotyping and detection. Subsequently, stained cells were conjugated with LIVE/DEAD Fixable Near-IR viability dye (Invitrogen, L34975) at 1:1000 dilution in PBS for 5 minutes at room temperature. Finally, cells were washed three times in cold cell media (RPMI, 10% FBS) before fluorescence-activated cell sorting (BD Biosciences, FACSAria Fusion). CAR+ sorting gates were drawn based on fluorescence of PBMCs from a similarly stained healthy donor.

Sorted endogenous immune cells (CAR^-^) and CAR T cells (CD3^+^CAR^+^) were separately partitioned into droplets for single-cell RNA-seq/CITE-seq/TCR-seq via Chromium Next GEM Single-Cell 5’Kit v2 (10x Genomics, 1000263). RNA-seq libraries were prepared according to manufacturer protocols. CITE-seq libraries were prepared via the 5’ Feature Barcode Kit (10x Genomics, 1000256). TCR-seq libraries were prepared via the Chromium Single-Cell Human TCR Amplification Kit (10x Genomics, 1000252). All libraries (RNA-seq, CITE-seq, TCR-seq) were quantified via the Qubit dsDNA HS Assay Kit (Invitrogen, Q32851), quality-checked for fragment sizes via high-sensitivity D5000 screentapes (Agilent, 5067-5592), pooled, and sequenced (Illumina, NovaSeq 6000 and NovaSeq X).

### CITE-seq antibody preparation

Thirty-seven human “Cellular Indexing of Transcriptomes and Epitopes by Sequencing” (CITE-seq^70^) antibodies were obtained from BioLegend (TotalSeq-C reagents): anti-CD3ε (clone UCHT1, 300479), anti-CD5 (clone UCHT2, 300637), anti-CD4 (clone SK3, 344651), anti-CD8α (clone SK1, 344753), anti-CD45RA (clone HI100, 304163), anti-CD45RO (clone UCHL1, 304259), anti-CCR7 (clone G043H7, 353251), anti-CD95 (clone DX2, 305651), anti-CD57 (clone QA17A04, 393321), anti-CD25 (clone BC96, 302649), anti-CD127 (clone A019D5, 351356), anti-CD103 (clone Ber-ACT8, 350233), anti-CXCR3 (clone G025H7, 353747), anti-CCR4 (clone L291H4, 359425), anti-CCR6 (clone G034E3, 353440), anti-PD-1 (clone EH12.2H7, 329963), anti-TIM-3 (clone F38-2E2, 345049), anti-LAG-3 (clone 11C3C65, 369335), anti-CD39 (clone A1, 328237), anti-TIGIT (clone A15153G, 372729), anti-CD27 (clone O323, 302853), anti-CD40L (clone 24-31, 310849), anti-GITR (clone 108-17, 371227), anti-OX40 (clone Ber-ACT35, 350035), anti-4-1BB (clone 4B4-1, 309839), anti-CD28 (clone CD28.2, 302963), anti-CD62L (clone DREG-56, 304851), anti-KLRG1 (clone SA231A2, 367737), anti-CD7 (clone CD7-6B7, 343127), anti-CD38 (clone HIT2, 303543), anti-HLA-DR (clone L243, 307663), anti-CD69 (clone FN50, 310951), anti-GARP (clone 7B11, 352517), anti-LAP (clone S20006A, 300015), anti-CXCR4 (clone 12G5, 306533), anti-CXCR5 (clone J252D4, 356939), anti-CXCR6 (clone K041E5, 356023), and anti-CX3CR1 (K0124E1, 355705). Residual infusion product cells were stained with all 37 CITE-seq antibodies. Patient peripheral blood mononuclear cells were stained with all CITE-seq antibodies minus phenotyping antibodies (described below). To prepare the staining solution, the antibody pool was constructed and centrifuged at 14000 × g for 10 minutes at room temperature in FACS buffer (PBS, 2% BSA, 0.05% sodium azide) to remove aggregates. The antibody supernatant was extracted and diluted with fresh FACS buffer to appropriate staining concentrations.

### Phenotyping antibody preparation

Antibodies for phenotyping T cells were obtained from BioLegend: anti-CD3ε (clone SK7, 344833), anti-CD4 (clone SK3, 344691), and anti-CD8α (clone SK1, 344716). To prepare the staining solution, antibodies were pooled in FACS buffer at 1:100 dilution together with CITE-seq antibodies.

### Single-cell sequencing data processing

Paired RNA-seq and CITE-seq reads were aligned with the Cell Ranger platform (version 7.1.0)^71^ to a custom GRCh38 reference genome modified to include the idecabtagene vicleucel and ciltacabtagene autoleucel CAR transgene sequences. TCR-seq reads were aligned with the Cell Ranger platform (version 7.1.0) to the GRCh38 vdj reference genome.

### Preprocessing, dimensionality reduction, and clustering

ScRNA-seq data preprocessing, dimensionality reduction, and clustering were performed using the Seurat package^72^ (version 5.0.0). Raw counts were log-normalized, and cells with >20% mitochondrial content or < 300 RNA features were removed. The top 10,000 variable genes were identified, and the data matrix was centered and scaled to regress out the effect of cell cycle signatures, percentage of mitochondrial RNA content, UMI count, and unique gene count. TCR genes were excluded from the list of variable genes. For analysis of cell subsets, a range of 2,000-8,000 variable genes were selected. Principal Component Analysis (PCA) and integration across patients and tissues were performed using the Harmony^73^ (version 1.2.0) package, and the top 50 harmony components were used for UMAP dimensionality reduction. The top 50 harmony components were then used to construct a Shared Nearest Neighbor (SNN) graph. Clusters were identified using the Louvain algorithm and annotated based on marker gene expression. CITE-seq data was centered log ratio (CLR) normalized across features.

### Differential gene expression (DEG) analysis, gene set enrichment analysis (GSEA), and gene set signature scoring

DEG analysis between groups of cells was performed using the Wilcoxon rank-sum test with Bonferroni correction. DEGs were filtered to only include genes with adjusted p-values < .05. GSEA was performed on DEGs using the ClusterProfiler package^74^ (version 4.6.2) and curated pathways from the MSigDB^75^ database. For GSEA on cytokine immune signatures^76^, cell type-specific (e.g. CD8^+^ T cell) pathways were used. Enriched pathways were filtered to only include pathways with FDR-adjusted q-values < 0.05. Gene set signature scores were calculated using the Seurat *AddModuleScore* function with default parameters.

### Gene expression visualization

Feature plots and violin plots depict normalized gene expression. Volcano plots depict log_2_fold-change in gene expression. Heatmaps were generated using the ComplexHeatmap package^77^ (version 2.14.0) and depict scaled average gene expression. All genes included in heatmaps contrasting treatments were found to be differentially expressed between treatments (adjusted p-values < .05) using the Wilcoxon rank-sum test with Bonferroni correction.

### Regulon analysis

Regulon analysis was performed using the pySCENIC package^78,79^ (version 0.12.0) multi-run workflow. Specifically, we first ran the recommended workflow 5 times and aggregated all the transcription factors (TFs) that appeared at least once into a filtered TF set. We then repeated the recommended workflow 100 times using the filtered TF set. We assigned each regulon an importance score based on the total number of times it appeared across all the runs. For each regulon, we assigned its target genes an importance score based on the number of times it was found in the regulon across all runs. We then calculated a regulon score (weighted by gene importance) for each cell using the AUCell package^79^ (version 1.20.2) on raw gene expression counts. Differential regulon score expression between groups of cells was performed using the Wilcoxon rank-sum test with Bonferroni correction.

### Single-cell TCR-seq analysis

Aggregation of TCR chains into clonotypes, clonotype overlap and tracing analysis, and clonal visualization were all performed using the scRepertoire package^80^ (version 2.2.1). TCR clonotypes were defined by the nucleotide sequence of the CDR3α and CDR3β regions and VDJC gene sequence. Clonotypes with only one TCR chain were removed, and clonotypes with more than two TCR chains were filtered to include only the top two expressed chains. TCR clonotypes were integrated with RNA expression based on matching barcodes. Large clones were defined as clonotypes that occupied greater than 1% of a sample’s TCR repertoire, medium clones were defined as clonotypes that occupied between 0.1-1%, and small clones were defined as clonotypes that occupied between 0.01-.01%.

### Gene module discovery

Module discovery was performed using the hdWGCNA package^81^ (version 0.3.01). Genes expressed by at least 5% of cells were kept, and metacells were generated within samples using Harmony embeddings without cell sharing. To construct the network, a minimum module size of 15 and a deep split parameter of 3 were chosen. The top 150 genes ranked by eigengene-based connectivity for each module were used to compute module scores.

### Single-cell copy number variation (CNV) analysis

Single-cell CNV analysis was performed using the numbat package^82^ (version 1.4.2) on cells annotated as B and plasma cells. Default settings were used aside from setting the *min_cells* parameter to 10.

### Ligand-receptor analysis

Ligand-receptor interaction analysis was performed using the LIANA package^83^ (version 0.1.13). We separately performed LIANA on all cell subsets for each bone marrow sample by setting *expr_prop* to 0.1 and *min_cells* to 10. To find differentially upregulated ligands and receptors in the post-ide-cel conventional monocytes subset, we first tested for DEGs between post-ide-cel and post-cilta-cel conventional monocytes. We then filtered the post-ide-cel conventional monocyte interactions to only include interactions that featured a post-ide-cel upregulated DEG ligand or receptor. We separately performed over-representation analysis on post-ide-cel upregulated plasma cell ligands and conventional monocyte receptors using the ClusterProfiler package and MSigDB pathways. Enriched pathways were filtered to only include pathways with FDR-adjusted q-values < 0.05.

### Gene trajectory analysis

Gene trajectory analysis was performed using the GeneTrajectory package^84^ (version 1.0.0) using default parameters.

### Data availability

All data are available from the authors upon reasonable request.

## Data Availability

All data produced in the present study are available upon reasonable request to the authors

## AUTHOR CONTRIBUTIONS

E.T., T.P., and J.H. conceived and designed all experiments. E.T. performed flow cytometry. E.T. and N. A. performed single-cell data generation. T.P. performed all single-cell analyses. T.P. and E.T. interpreted flow cytometry and single-cell data. J.H. supervised the project. Y.H. and B.D. collected and interpreted clinical data. M.S., T.A., and P.A.R. coordinated patient biospecimens. T.P. and J.H. prepared the manuscript. All authors reviewed the manuscript.

## ACKNOWLEDGEMENTS

This work was supported by the NIH R21AI169159 grant and American Cancer Society Scholar Award (SG-22-136-01-1BCD) (to J.H.). T.P. was supported by the University of Chicago CSTR training grant (T32HL007381). Y.H. was supported by the University of Chicago MSTP Training Grant (T32GM007281). N.A. was supported by the University of Chicago MTCR training grant (T32CA009594). We thank the UChicago Human Immunologic Monitoring Facility for patient biospecimen cryopreservation, as well as the Pritzker School of Molecular Engineering Single-Cell Immunophenotyping Core and UChicago Genomics Facility for assisting in the generation of single-cell sequencing data. This work was completed in part with resources provided by the University of Chicago’s Research Computing Center.

