## Supplementary Data for "Cilta-cel salvages ide-cel failure in relapsed multiple myeloma by driving distinct immune responses"

**Tables and supplemental figures for:**

| **Diagnosis** |
| --- |
| IgG-lambda myeloma |
| FGFR3/IGH t(4;14) rearrangement |
| Monosomy 13 |

**Supplemental table 1: Clinical characteristics of the patient**

| Line | Therapy | Dose | Response | Notes |
|---|---|---|---|---|
| | Lenalidomide<br>Bortezomib<br>Dexamethasone | 4 cycles | VGPR | |
| 1 | Tandem autologous stem cell transplant<br>Lenalidomide | $4.6 \times 10^6$ cells/kg x 2 doses | CR, MRD- | Relapse ~2 years later |
| 2 | Daratumumab<br>Carfilzomib<br>Dexamethasone | 4 cycles | PR | |
| 3 | Fludarabine<br>Cyclophosphamide<br>Idecabtagene vicleucel<br>Pomalidomide<br>Dexamethasone<br>Cyclophosphamide | 300-460 $\times 10^6$ cells/kg | NR, PD | Grade 2 CRS |
| 4 | Cyclophosphamide<br>Pomalidomide<br>Dexamethasone | 1 cycle | | |
| 5 | Bendamustine<br>Ciltacabtagene autoleucel<br>Pomalidomide | $0.2 \times 10^6$ cells/kg | CR, MRD- | Grade 1 CRS |

### Supplemental table 2: Treatment history of the patient.

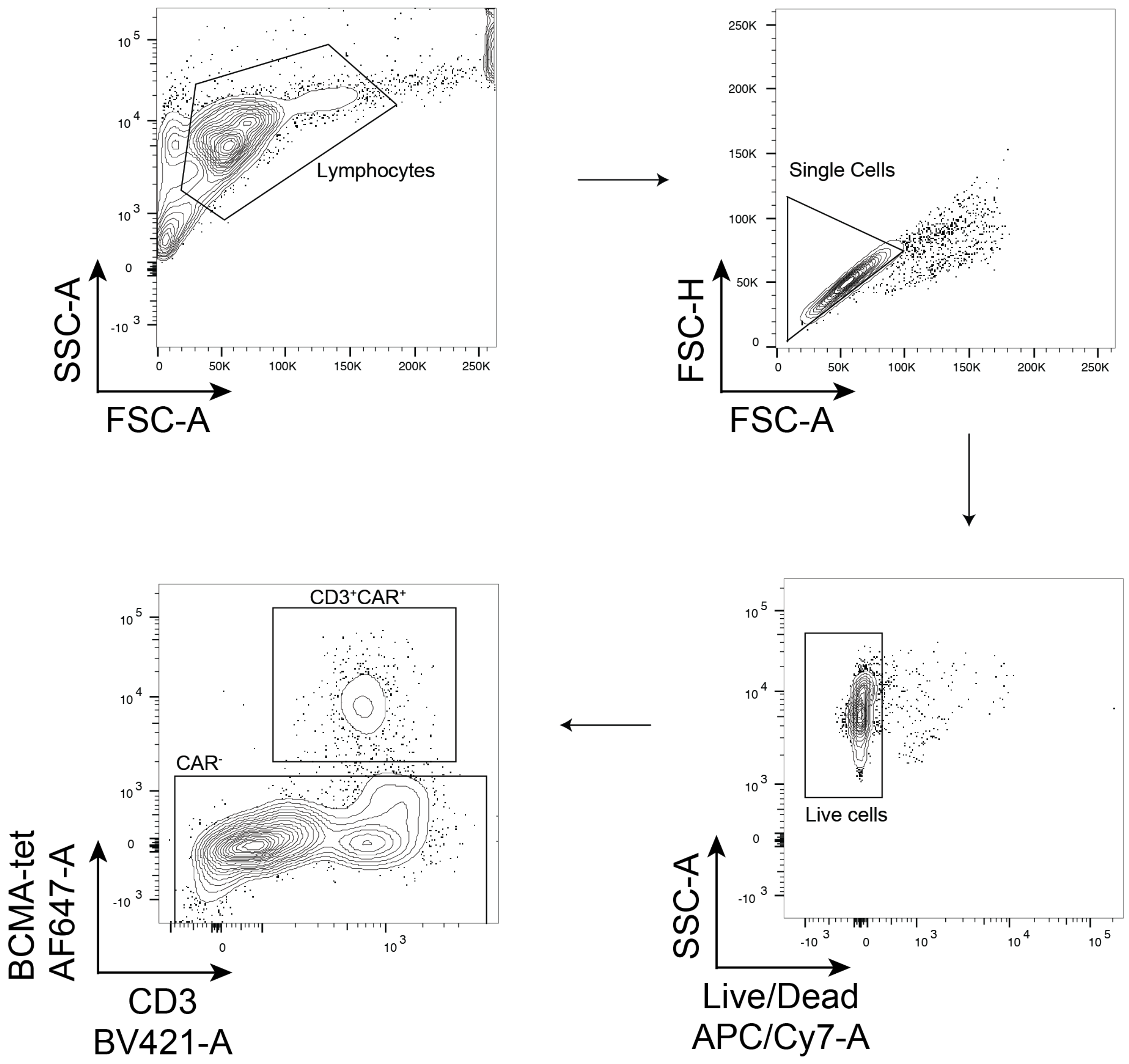


**Figure S1: CAR T-cell sorting and phenotyping strategy.** Representative flow plots demonstrating sorting strategy. $CD3^+CAR^+$ CAR T cells and $CAR^-$ cells were sorted. PBMCs from a healthy donor were used to draw $CAR^+$ gates.

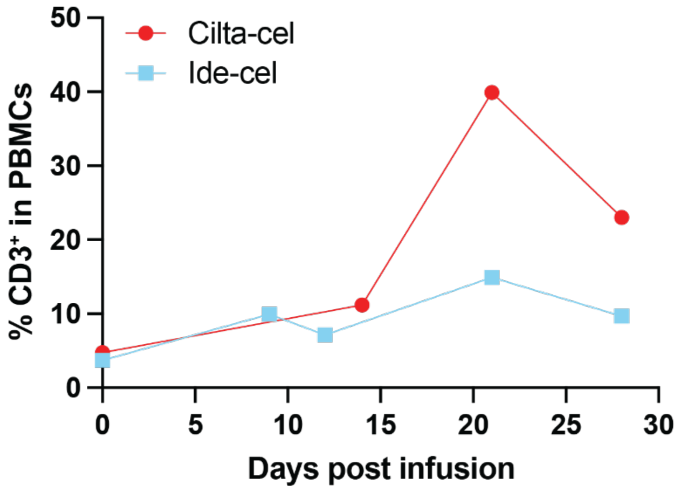


**Figure S2: Quantification of post-infusion CAR T cells in the peripheral blood.** Line plots depicting the total peripheral T cell ($CD3^+$) abundance at each timepoint, for each treatment, as determined by flow cytometry.

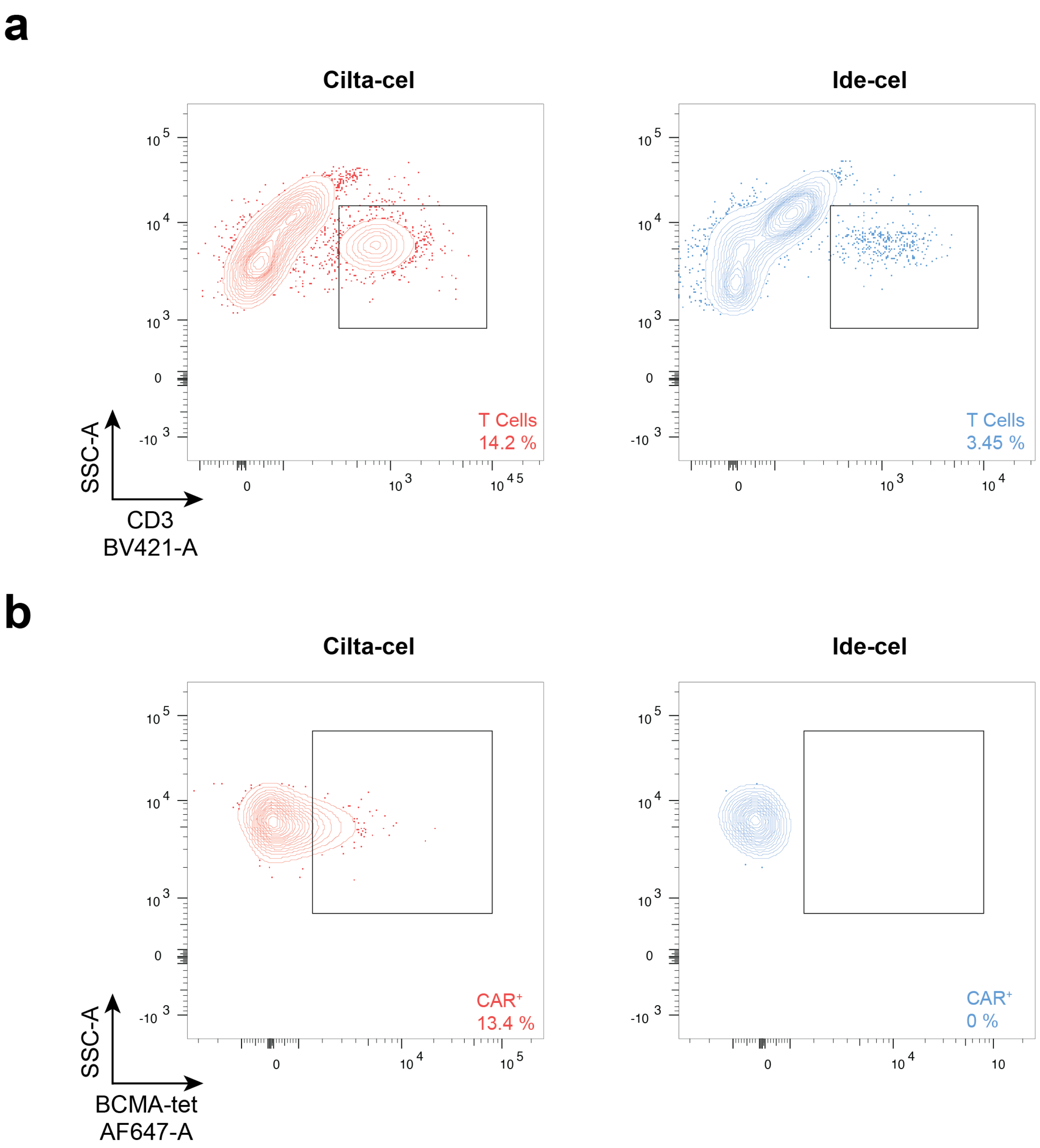


**Figure S3: Quantification of post-infusion CAR T cells in the bone marrow.** (**a**) Flow plots depicting post-infusion bone marrow CD3$^+$ T-cell abundance for each sample. (**b**) Flow plots depicting the post-infusion bone marrow CAR T-cell abundance for each sample. PBMCs from a healthy donor were used to draw CAR$^+$ gates.

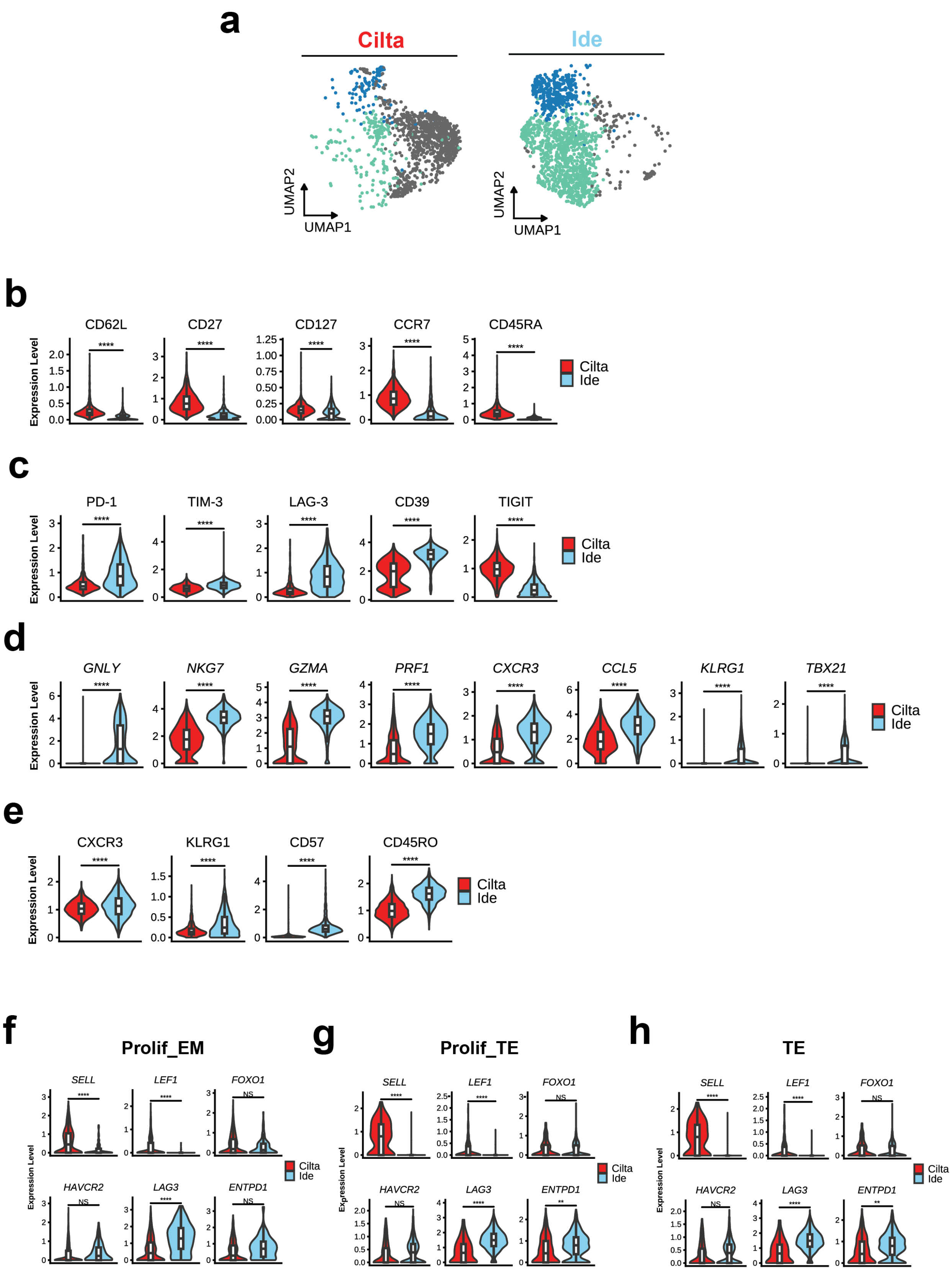
a
Cilta
Ide
UMAP2
UMAP1
b
CD62L
CD27
CD127
CCR7
CD45RA
Expression Level
Cilta
Ide
c
PD-1
TIM-3
LAG-3
CD39
TIGIT
d
GNLY
NKG7
GZMA
PRF1
CXCR3
CCL5
KLRG1
TBX21
e
CXCR3
KLRG1
CD57
CD45RO
f
Prolif_EM
g
Prolif_TE
h
TE
SELL
LEF1
FOXO1
HAVCR2
LAG3
ENTPD1
****
**
NS

**Figure S4: CD8$^+$ CAR T-cell infusion product phenotype proportions and gene and protein expression.** (**a**) UMAP plot of CD8$^+$ CAR T cells, colored by subset and split by treatment. (**b-e**) Violin plots contrasting expression of (**b**) proteins associated with T-cell memory, (**c**) T-cell inhibitory receptors, (**d**) genes associated with T-cell effector function and mature differentiation, and (**e**) proteins associated with T-cell effector function. Expression levels were compared by Wilcoxon rank-sum test with Bonferroni adjustment, whereby **** indicates p<.0001, ** indicates p<.01, and ns indicates not significant. (**f-g**) Violin plots contrasting memory-associated (top) and inhibitory receptor (bottom) gene expression of each treatment in the (**f**) proliferating effector memory, (**g**) proliferating terminal effector, and (**h**) terminal effector subsets. Expression levels were compared by Wilcoxon rank-sum test with Bonferroni adjustment, whereby **** indicates p<.0001 and ns indicates not significant.

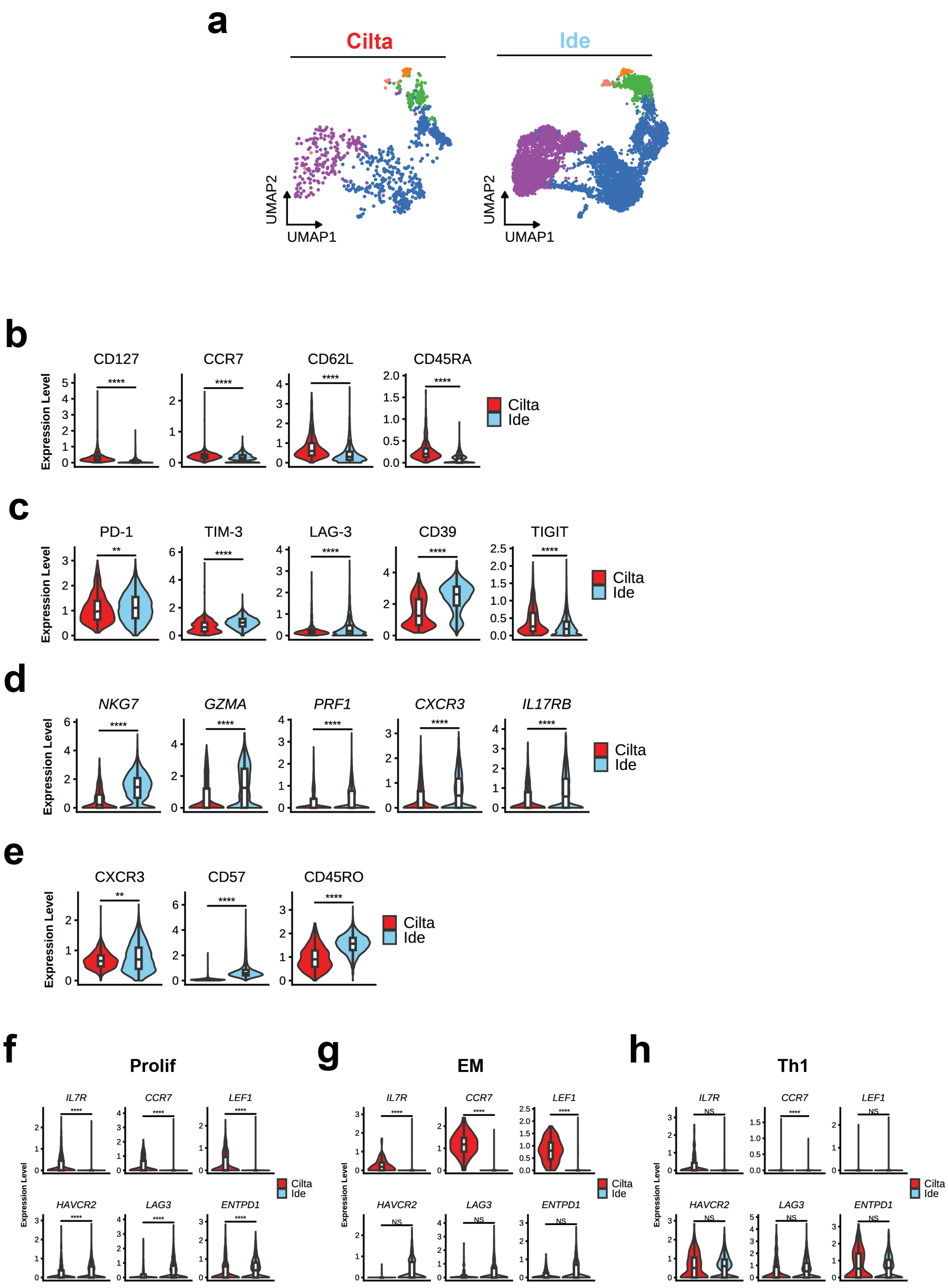
a
Cilta
Ide
UMAP2
UMAP1
b
CD127
CCR7
CD62L
CD45RA
Expression Level
Cilta
Ide
c
PD-1
TIM-3
LAG-3
CD39
TIGIT
d
NKG7
GZMA
PRF1
CXCR3
IL17RB
e
CXCR3
CD57
CD45RO
f
Prolif
g
EM
h
Th1
IL7R
CCR7
LEF1
HAVCR2
LAG3
ENTPD1
NS

**Figure S5: CD4+ CAR T-cell infusion product phenotype proportions and gene and protein expression. (a)** UMAP plot of CD4+ CAR T cells, colored by subset split and by treatment. (**b-e**) Violin plots contrasting expression of (**b**) proteins associated with T-cell memory, (**c**) T-cell inhibitory receptors, (**d**) genes associated with T-cell effector function and mature differentiation, and (**e**) proteins associated with T-cell effector function. Expression levels were compared by Wilcoxon rank-sum test with Bonferroni adjustment, whereby **** indicates p<.0001, ** indicates p<.01, and ns indicates not significant. (**f-g**) Violin plots contrasting memory-associated (top) and inhibitory receptor (bottom) gene expression of each treatment in the (**f**) proliferating, (**g**) effector memory, and (**h**) T helper type 1 subsets. Expression levels were compared by Wilcoxon rank-sum test with Bonferroni adjustment, whereby **** indicates p<.0001 and ns indicates not significant.

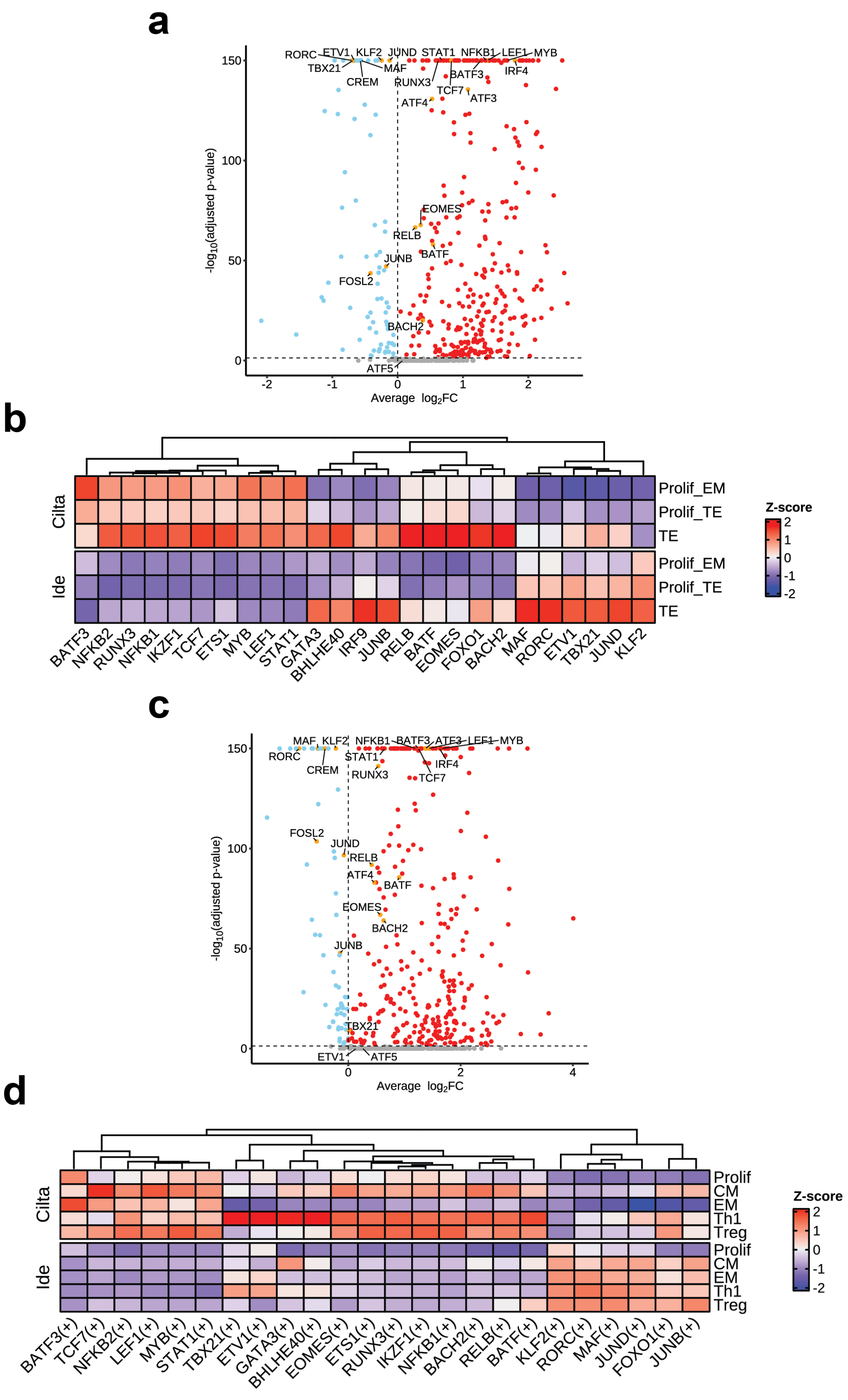

a
RORC
ETV1
KLF2
JUND
STAT1
NFKB1
LEF1
MYB
TBX21
MAF
CREM
RUNX3
BATF3
IRF4
TCF7
ATF3
ATF4
EOMES
RELB
BATF
JUNB
FOSL2
BACH2
ATF5
150
100
50
0
-log10(adjusted p-value)
-2
-1
0
1
2
Average log2FC
b
Cilta
Ide
Prolif_EM
Prolif_TE
TE
Prolif_EM
Prolif_TE
TE
Z-score
2
1
0
-1
-2
BATF3
NFKB2
RUNX3
NFKB1
IKZF1
TCF7
ETS1
MYB
LEF1
STAT1
GATA3
BHLHE40
IRF9
JUNB
RELB
BATF
EOMES
FOXO1
BACH2
MAF
RORC
ETV1
TBX21
JUND
KLF2
c
MAF
KLF2
NFKB1
BATF3
ATF3
LEF1
MYB
RORC
STAT1
CREM
IRF4
RUNX3
TCF7
FOSL2
JUND
RELB
ATF4
BATF
EOMES
BACH2
JUNB
TBX21
ETV1
ATF5
150
100
50
0
-log10(adjusted p-value)
0
2
4
Average log2FC
d
Cilta
Ide
Prolif
CM
EM
Th1
Treg
Prolif
CM
EM
Th1
Treg
Z-score
2
1
0
-1
-2
BATF3(+)
TCF7(+)
NFKB2(+)
LEF1(+)
MYB(+)
STAT1(+)
TBX21(+)
ETV1(+)
GATA3(+)
BHLHE40(+)
EOMES(+)
ETS1(+)
RUNX3(+)
IKZF1(+)
NFKB1(+)
BACH2(+)
RELB(+)
BATF(+)
KLF2(+)
RORC(+)
MAF(+)
JUND(+)
FOXO1(+)
JUNB(+)

**Figure S6: CAR T-cell infusion product regulon expression.** (**a**) Volcano plot depicting differentially expressed regulons by each treatment in $CD8^+$ CAR T cells. Significant regulons are colored according to $log_2$fold-change between expression in cilta-cel (red) and ide-cel (blue). Highlighted genes are colored in orange. (**b**) Heatmap depicting scaled expression of selected differentially expressed regulons grouped by $CD8^+$ subset and split by treatment. Regulons are clustered by Euclidean distance. (**c**) as in (**a**), but for $CD4^+$ CAR T cells. (**d**) as in (**b**), but for $CD4^+$ CAR T cells.

**a**

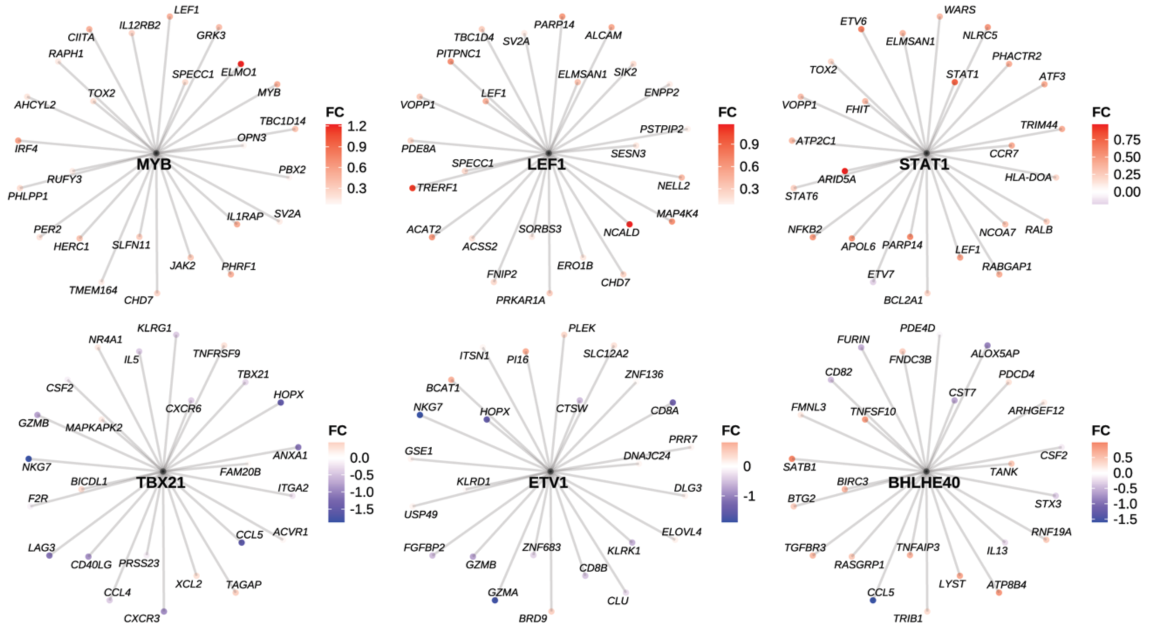
MYB
LEF1
IL12RB2
CIITA
GRK3
RAPH1
SPECC1
ELMO1
TOX2
MYB
AHCYL2
TBC1D14
OPN3
IRF4
PBX2
RUFY3
PHLPP1
IL1RAP
SV2A
PER2
HERC1
SLFN11
JAK2
PHRF1
TMEM164
CHD7
FC
1.2
0.9
0.6
0.3
LEF1
PARP14
TBC1D4
SV2A
ALCAM
PITPNC1
ELMSAN1
SIK2
LEF1
ENPP2
VOPP1
PSTPIP2
PDE8A
SESN3
SPECC1
TRERF1
NELL2
MAP4K4
ACAT2
SORBS3
NCALD
ACSS2
FNIP2
ERO1B
CHD7
PRKAR1A
FC
0.9
0.6
0.3
STAT1
WARS
ETV6
ELMSAN1
NLRC5
TOX2
PHACTR2
STAT1
ATF3
VOPP1
FHIT
TRIM44
ATP2C1
CCR7
ARID5A
HLA-DOA
STAT6
NFKB2
NCOA7
RALB
APOL6
PARP14
LEF1
ETV7
RABGAP1
BCL2A1
FC
0.75
0.50
0.25
0.00
TBX21
KLRG1
NR4A1
IL5
TNFRSF9
TBX21
CSF2
HOPX
CXCR6
GZMB
MAPKAPK2
ANXA1
NKG7
BICDL1
FAM20B
ITGA2
F2R
CCL5
ACVR1
LAG3
CD40LG
PRSS23
XCL2
TAGAP
CCL4
CXCR3
FC
0.0
-0.5
-1.0
-1.5
ETV1
PLEK
ITSN1
PI16
SLC12A2
ZNF136
BCAT1
CTSW
CD8A
NKG7
HOPX
PRR7
GSE1
DNAJC24
KLRD1
DLG3
USP49
ELOVL4
FGFBP2
ZNF683
KLRK1
GZMB
CD8B
GZMA
CLU
BRD9
FC
0
-1
BHLHE40
PDE4D
FURIN
FNDC3B
ALOX5AP
CD82
PDCD4
CST7
FMNL3
TNFSF10
ARHGEF12
CSF2
SATB1
TANK
BIRC3
BTG2
STX3
RNF19A
TGFBR3
TNFAIP3
IL13
RASGRP1
LYST
ATP8B4
CCL5
TRIB1
FC
0.5
0.0
-0.5
-1.0
-1.5

**b**

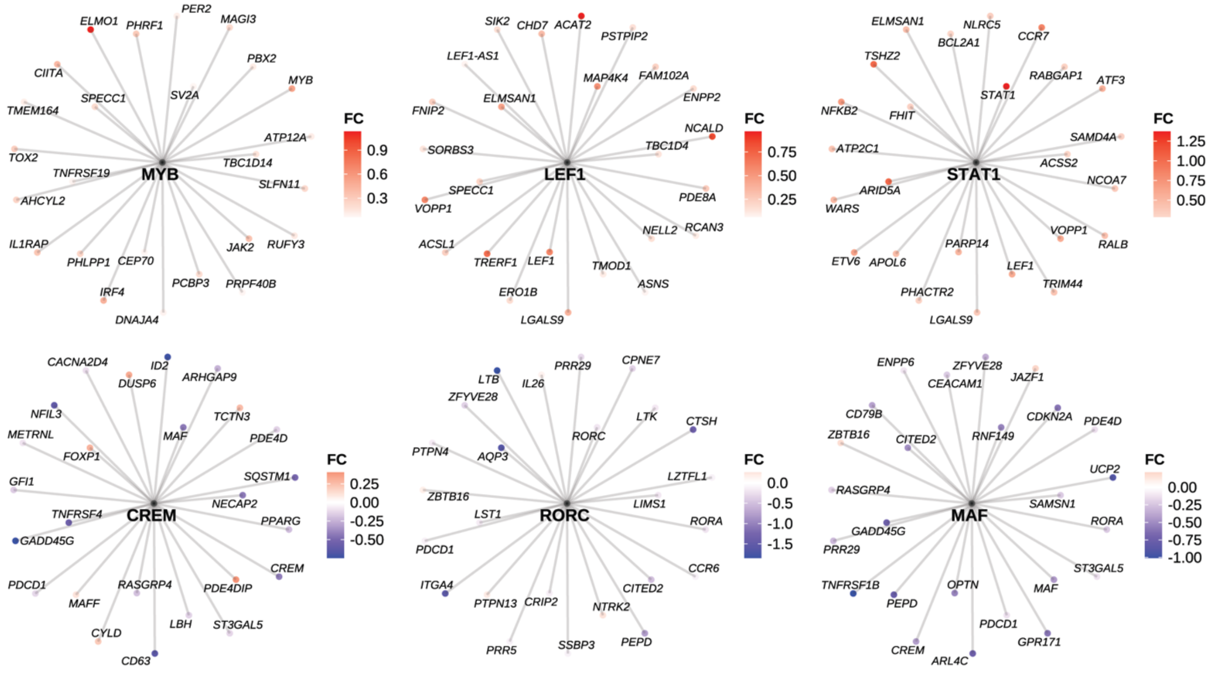
MYB
PER2
ELMO1
PHRF1
MAGI3
PBX2
CIITA
MYB
SPECC1
SV2A
TMEM164
ATP12A
TOX2
TBC1D14
TNFRSF19
SLFN11
AHCYL2
IL1RAP
JAK2
RUFY3
PHLPP1
CEP70
PCBP3
PRPF40B
IRF4
DNAJA4
FC
0.9
0.6
0.3
LEF1
SIK2
CHD7
ACAT2
PSTPIP2
LEF1-AS1
MAP4K4
FAM102A
ELMSAN1
ENPP2
FNIP2
NCALD
TBC1D4
SORBS3
SPECC1
PDE8A
VOPP1
NELL2
RCAN3
ACSL1
TRERF1
LEF1
TMOD1
ASNS
ERO1B
LGALS9
FC
0.75
0.50
0.25
STAT1
ELMSAN1
NLRC5
BCL2A1
CCR7
TSHZ2
RABGAP1
ATF3
STAT1
NFKB2
FHIT
SAMD4A
ATP2C1
ACSS2
NCOA7
ARID5A
WARS
VOPP1
RALB
PARP14
ETV6
APOL6
LEF1
TRIM44
PHACTR2
LGALS9
FC
1.25
1.00
0.75
0.50
CREM
CACNA2D4
ID2
DUSP6
ARHGAP9
NFIL3
TCTN3
METRNL
MAF
PDE4D
FOXP1
SQSTM1
GFI1
NECAP2
TNFRSF4
PPARG
GADD45G
CREM
PDCD1
RASGRP4
PDE4DIP
MAFF
LBH
ST3GAL5
CYLD
CD63
FC
0.25
0.00
-0.25
-0.50
RORC
CPNE7
PRR29
LTB
IL26
ZFYVE28
LTK
CTSH
RORC
PTPN4
AQP3
LZTFL1
ZBTB16
LIMS1
LST1
RORA
PDCD1
CCR6
ITGA4
CITED2
PTPN13
CRIP2
NTRK2
PEPD
PRR5
SSBP3
FC
0.0
-0.5
-1.0
-1.5
MAF
ENPP6
ZFYVE28
CEACAM1
JAZF1
CD79B
CDKN2A
PDE4D
ZBTB16
CITED2
RNF149
UCP2
RASGRP4
SAMSN1
GADD45G
RORA
PRR29
ST3GAL5
TNFRSF1B
OPTN
MAF
PEPE
PDCD1
GPR171
CREM
ARL4C
FC
0.00
-0.25
-0.50
-0.75
-1.00

**Figure S7: CAR T cell regulon membership.** (**a**) Network plots of selected treatment-associated regulons and their target genes in $CD8^+$ CAR T cells. In each regulatory network, only the top weighted genes are depicted. Genes are colored according to $\log_2$fold-change between expression in cilta-cel (red) and ide-cel (blue) $CD8^+$ CAR T cells. (**b**) As in (**a**), but for $CD4^+$ CAR T cells.

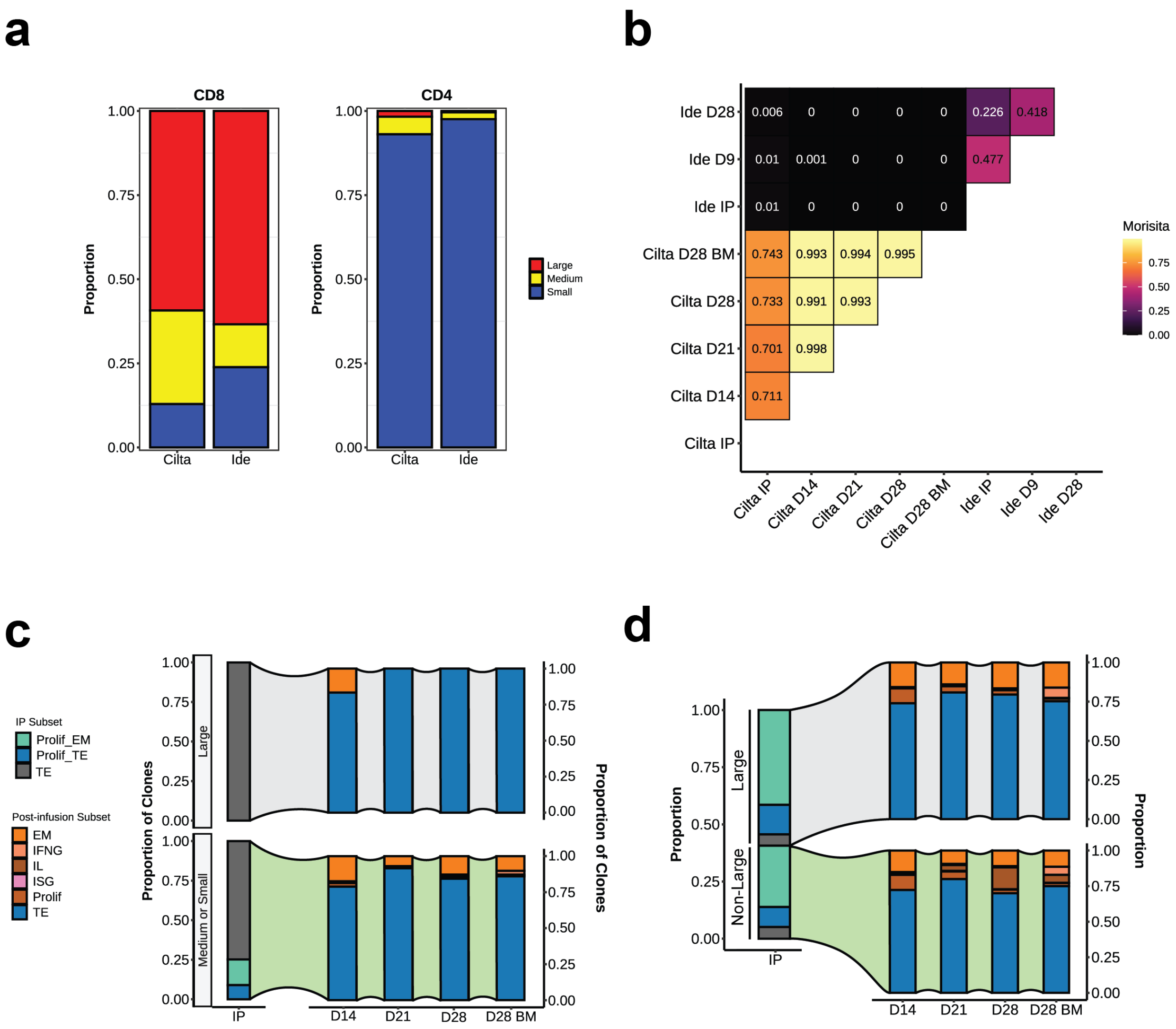
a
b
c
d
CD8
CD4
Proportion
Cilta
Ide
Large
Medium
Small
Ide D28
Ide D9
Ide IP
Cilta D28 BM
Cilta D28
Cilta D21
Cilta D14
Cilta IP
0.006
0.01
0.001
0.226
0.418
0.477
0.743
0.993
0.994
0.995
0.733
0.991
0.993
0.701
0.998
0.711
Morisita
IP Subset
Prolif_EM
Prolif_TE
TE
Post-infusion Subset
EM
IFNG
IL
ISG
Prolif
TE
Large
Medium or Small
Proportion of Clones
IP
D14
D21
D28
D28 BM
Non-Large

**Figure S8: CAR T-cell clonal repertoire overlap and phenotypic analysis.** (**a**) Barplots depicting the proportion of large, medium, and small clones in $CD8^+$ (left) and $CD4^+$ (right) infusion product CAR T cells for each treatment. (**b**) Heatmap depicting the Morisita overlap coefficients between each CAR T-cell sample. (**c**) Bar plots depicting the predominant phenotype for each cilta-cel $CD8^+$ CAR clonotypes that was present in both the infusion product and post-infusion, aggregated across large (top) versus non-large (bottom) clones. (**d**) As in (**c**) but depicting the proportion of each $CD8^+$ phenotype associated with large (top) versus non-large cilta-cel clonotypes (bottom).

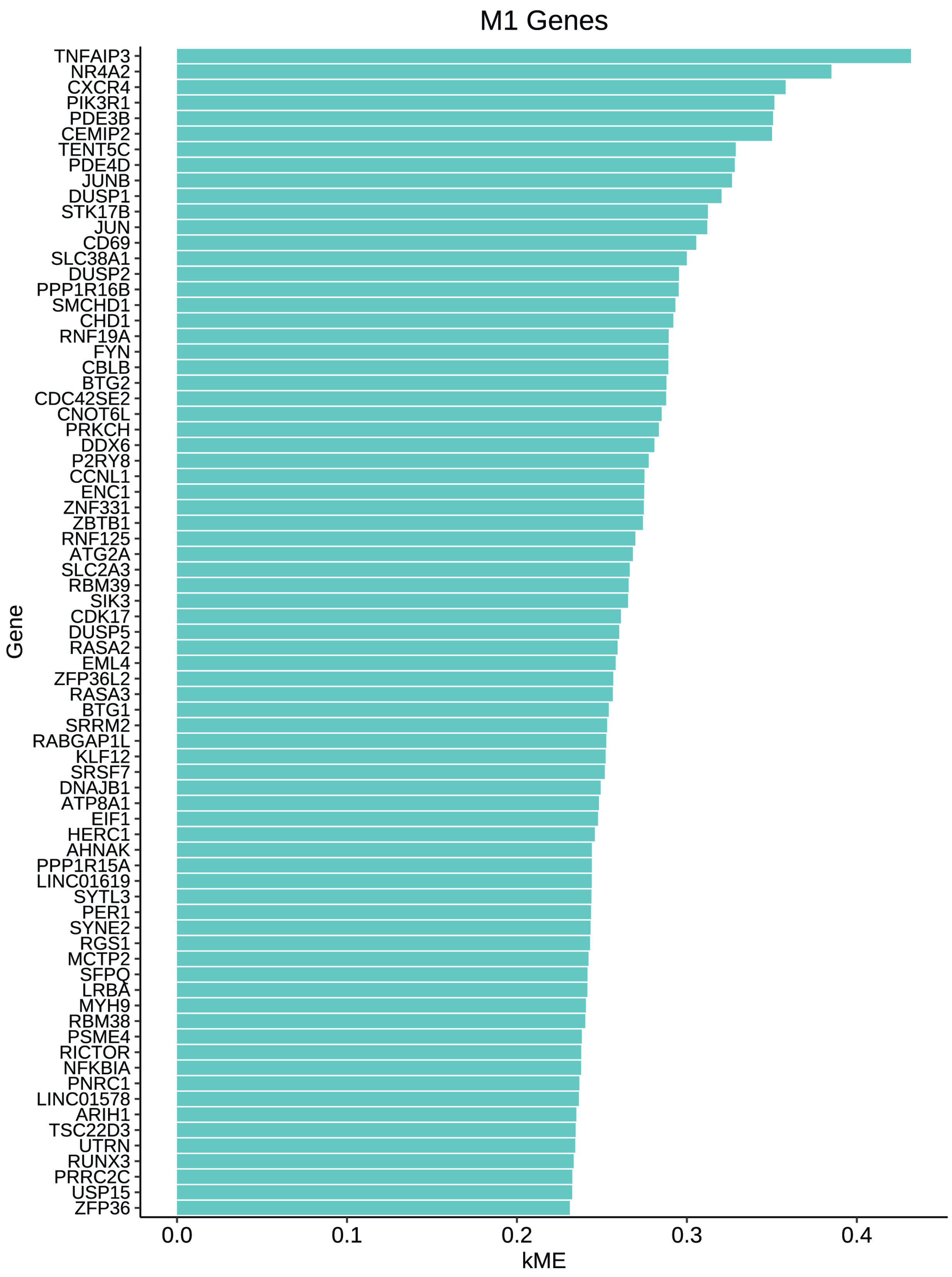
M1 Genes
TNFAIP3
NR4A2
CXCR4
PIK3R1
PDE3B
CEMIP2
TENT5C
PDE4D
JUNB
DUSP1
STK17B
JUN
CD69
SLC38A1
DUSP2
PPP1R16B
SMCHD1
CHD1
RNF19A
FYN
CBLB
BTG2
CDC42SE2
CNOT6L
PRKCH
DDX6
P2RY8
CCNL1
ENC1
ZNF331
ZBTB1
RNF125
ATG2A
SLC2A3
RBM39
SIK3
CDK17
DUSP5
RASA2
EML4
ZFP36L2
RASA3
BTG1
SRRM2
RABGAP1L
KLF12
SRSF7
DNAJB1
ATP8A1
EIF1
HERC1
AHNAK
PPP1R15A
LINC01619
SYTL3
PER1
SYNE2
RGS1
MCTP2
SFPQ
LRBA
MYH9
RBM38
PSME4
RICTOR
NFKBIA
PNRC1
LINC01578
ARIH1
TSC22D3
UTRN
RUNX3
PRRC2C
USP15
ZFP36
Gene
0.0
0.1
0.2
0.3
0.4
kME

**Figure S9: Post-infusion cilta-cel CD8$^+$ TE CAR T-cell M1 gene module membership.** Bar plots depicting the top 75 genes in module M1. Genes are ranked by eigengene-based connectivity (kME).

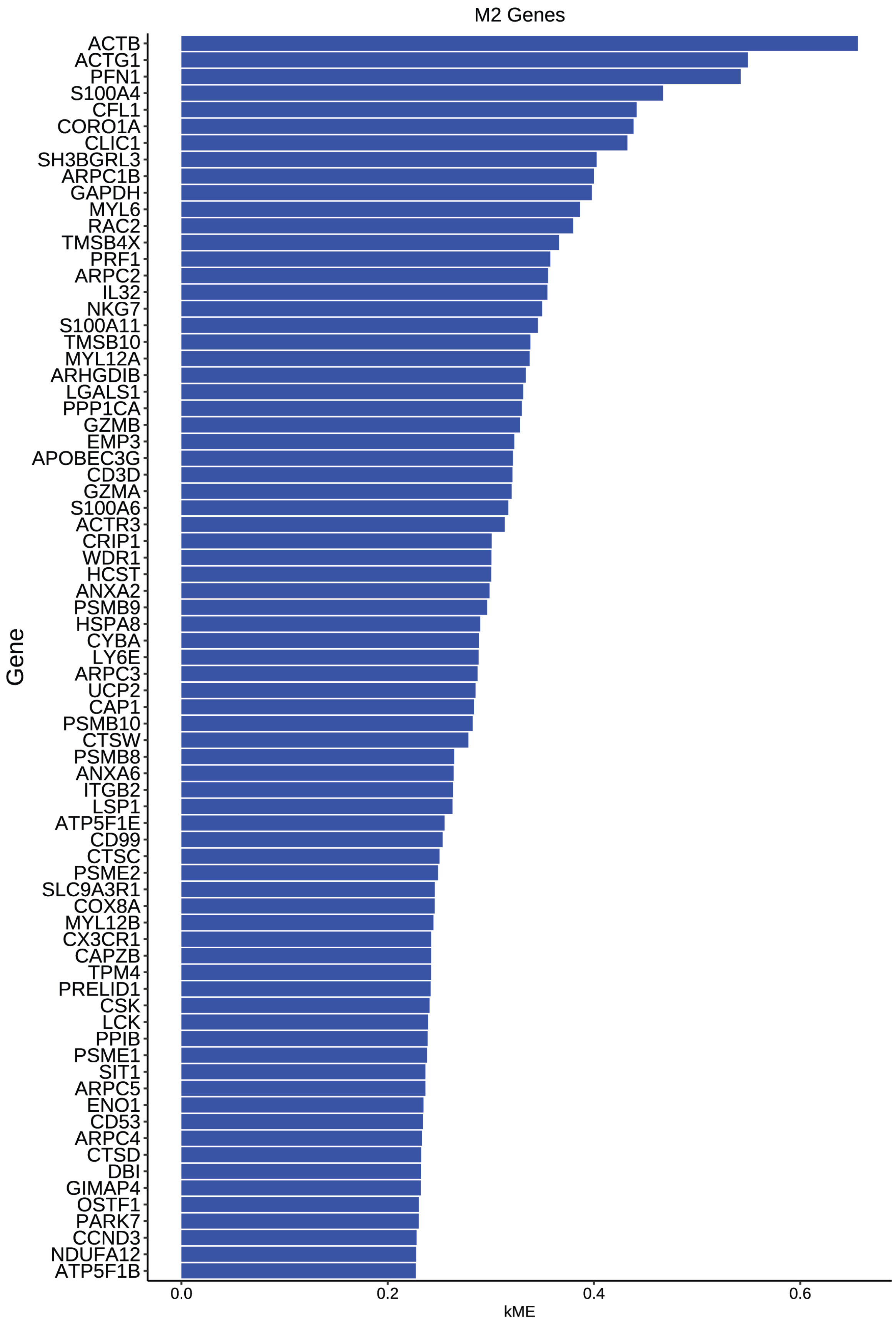
M2 Genes
Gene
kME
0.0
0.2
0.4
0.6
ACTB
ACTG1
PFN1
S100A4
CFL1
CORO1A
CLIC1
SH3BGRL3
ARPC1B
GAPDH
MYL6
RAC2
TMSB4X
PRF1
ARPC2
IL32
NKG7
S100A11
TMSB10
MYL12A
ARHGDIB
LGALS1
PPP1CA
GZMB
EMP3
APOBEC3G
CD3D
GZMA
S100A6
ACTR3
CRIP1
WDR1
HCST
ANXA2
PSMB9
HSPA8
CYBA
LY6E
ARPC3
UCP2
CAP1
PSMB10
CTSW
PSMB8
ANXA6
ITGB2
LSP1
ATP5F1E
CD99
CTSC
PSME2
SLC9A3R1
COX8A
MYL12B
CX3CR1
CAPZB
TPM4
PRELID1
CSK
LCK
PPIB
PSME1
SIT1
ARPC5
ENO1
CD53
ARPC4
CTSD
DBI
GIMAP4
OSTF1
PARK7
CCND3
NDUFA12
ATP5F1B

**Figure S10: Post-infusion cilta-cel CD8$^+$ TE CAR T-cell M2 gene module membership.** Bar plots depicting the top 75 genes in module M2. Genes are ranked by eigengene-based connectivity (kME).

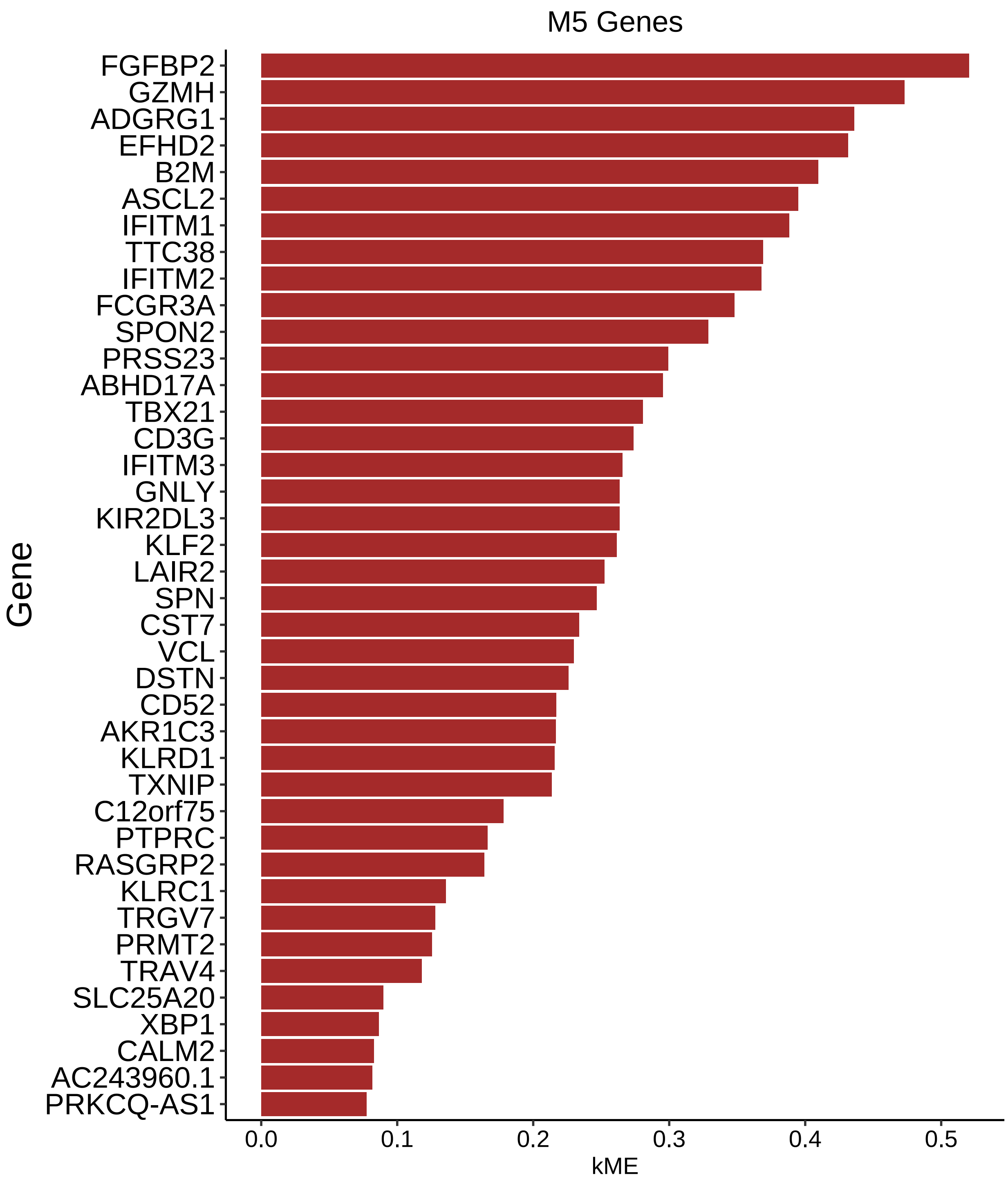


**Figure S11: Post-infusion cilta-cel CD8$^{+}$ TE CAR T-cell M5 gene module membership.** Bar plots depicting the genes in module M5. Genes are ranked by eigengene-based connectivity (kME).

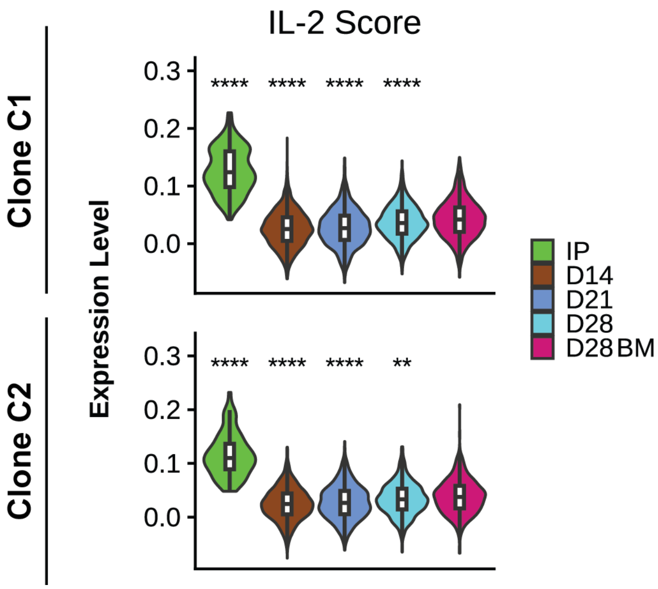


**Figure S12: Expression of the IL-2 gene signature at each timepoint of cilta-cel CAR T cells.** Violin plots depicting the expression of the IL-2 gene signature scores at each timepoint for clone C1 (top) and C2 (bottom). All significance comparisons are made relative to the D28 BM timepoint using the using the Wilcoxon rank-sum test with Bonferroni adjustment, whereby **** indicates p<.0001 and ** indicates p<.01.

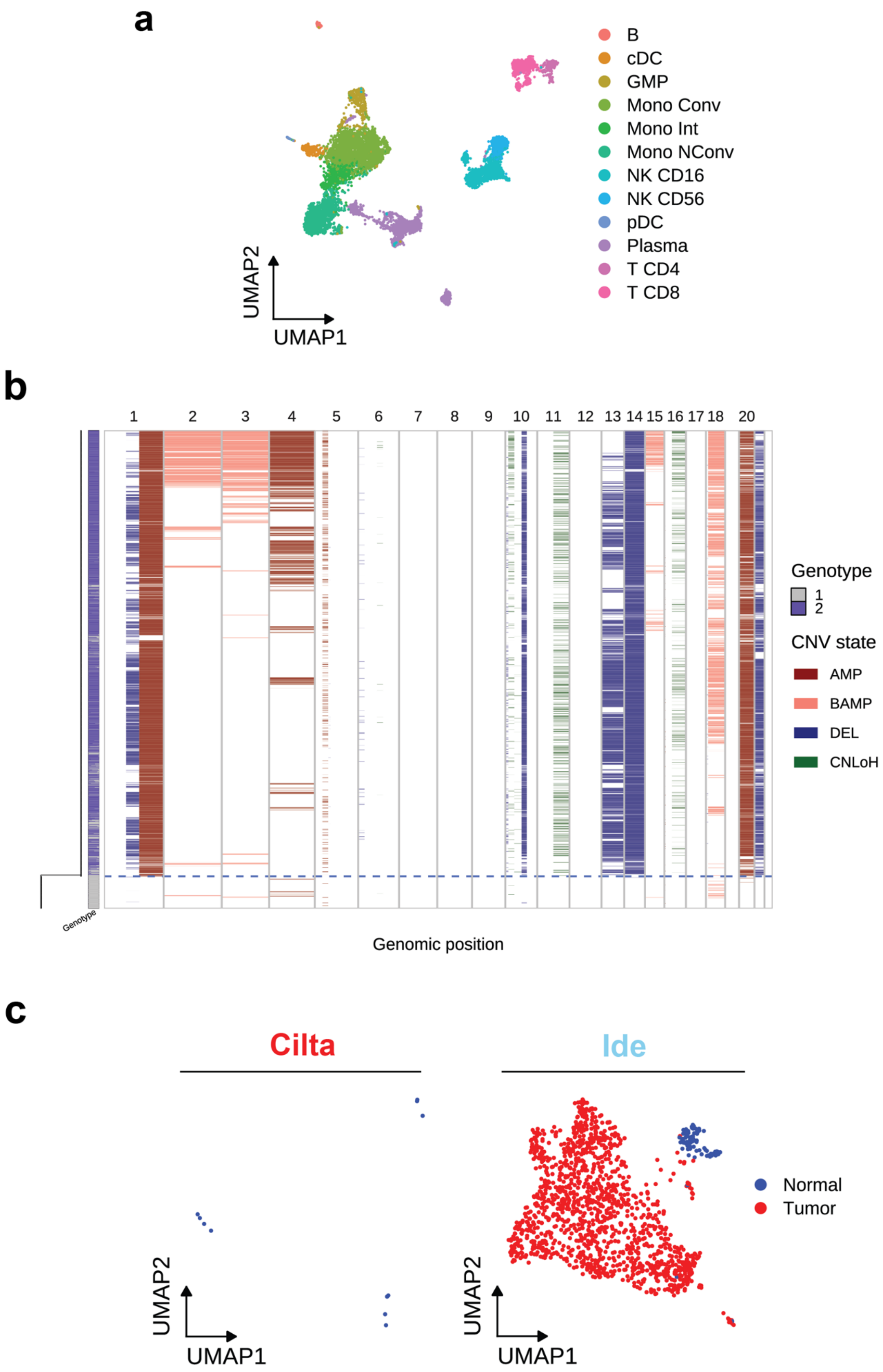
a
B
cDC
GMP
Mono Conv
Mono Int
Mono NConv
NK CD16
NK CD56
pDC
Plasma
T CD4
T CD8
UMAP2
UMAP1
b
1 2 3 4 5 6 7 8 9 10 11 12 13 14 15 16 17 18 20
Genotype
1
2
CNV state
AMP
BAMP
DEL
CNLoH
Genotype
Genomic position
c
Cilta
Ide
Normal
Tumor
UMAP2
UMAP1
UMAP2
UMAP1

**Figure S13: Phenotypic and characteristics of post-infusion endogenous immune and myeloma cells in the bone marrow.** (**a**) UMAP plot of the endogenous immune populations found in the bone marrow following CAR T-cell infusion, colored by subset. (**b**) Single-cell CNV landscape of bone marrow myeloma cells. Blue dashed line separates predicted tumor (genotype 1) and normal (genotype 2) cells. (**c**) UMAP plot of all B-lineage cells, colored by predicted tumor status and split by treatment. Abbreviations: CNV, copy number variation; AMP, amplification; BAMP, balanced amplification; DEL, deletion; CNLoH, copy-neutral loss of heterozygosity.

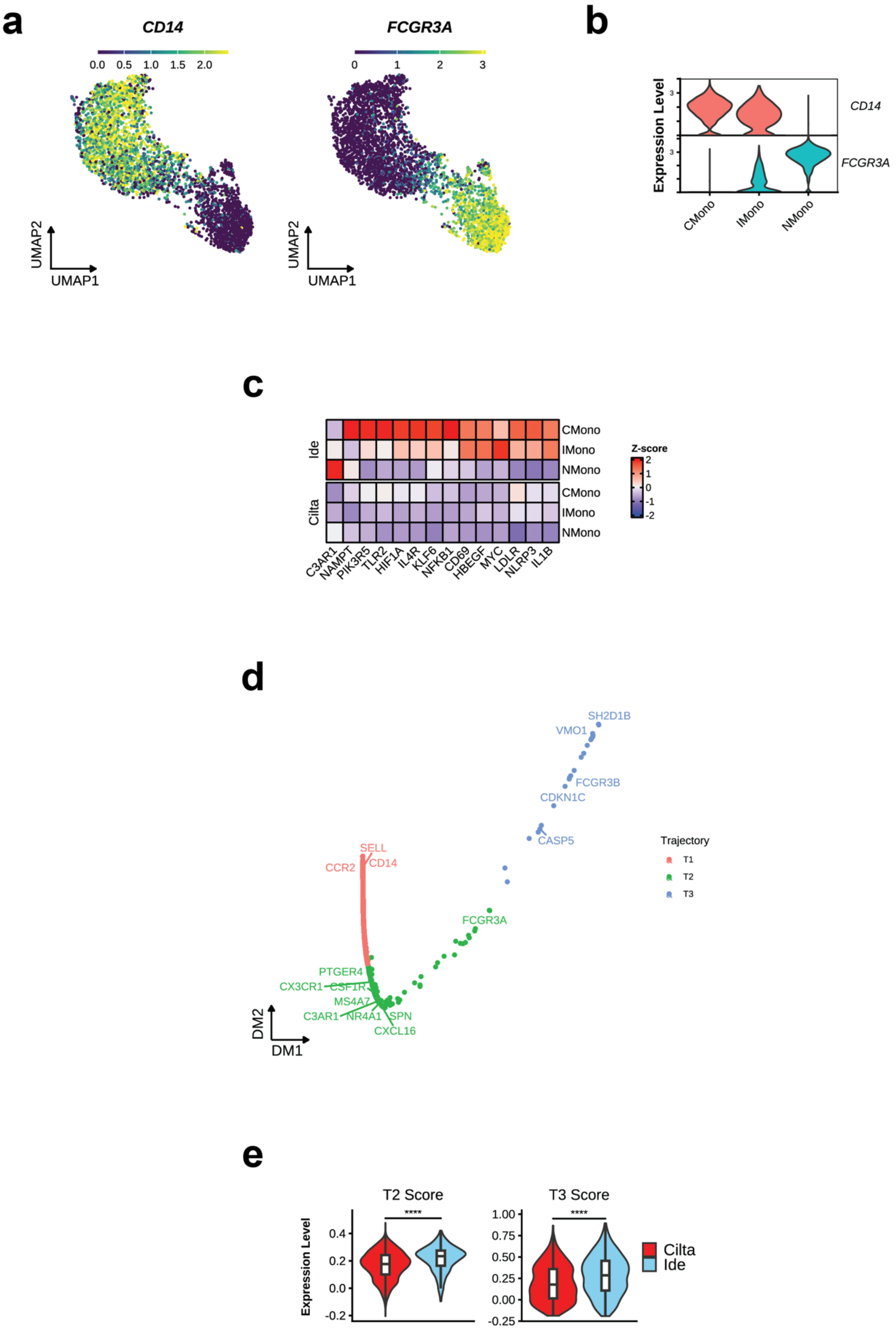
a
CD14
0.0 0.5 1.0 1.5 2.0
FCGR3A
0 1 2 3
UMAP2
UMAP1
UMAP2
UMAP1
b
Expression Level
CD14
FCGR3A
CMono
IMono
NMono
c
Ide
Cilta
CMono
IMono
NMono
CMono
IMono
NMono
Z-score
2
1
0
-1
-2
C3AR1
NAMPT
PIK3R5
TLR2
HIF1A
IL4R
KLF6
NFKB1
CD69
HBEGF
MYC
LDLR
NLRP3
IL1B
d
SH2D1B
VMO1
FCGR3B
CDKN1C
CASP5
SELL
CCR2
CD14
FCGR3A
PTGER4
CX3CR1
CSF1R
MS4A7
C3AR1
NR4A1
SPN
CXCL16
Trajectory
T1
T2
T3
DM2
DM1
e
T2 Score
T3 Score
****
****
Expression Level
0.4
0.2
0.0
-0.2
1.00
0.75
0.50
0.25
0.00
-0.25
Cilta
Ide

**Figure S14: Phenotypic and transcriptomic characteristics of post-infusion monocytes cells in the bone marrow.** (**a**) Expression plots of the *CD14* and *FCGR3A* genes. (**b**) Violin plots of *CD14* and *FCGR3A* genes across phenotypes. (**c**) Heatmap depicting scaled expression of selected inflammatory DEGs in monocytes, grouped by phenotype and split by treatment. (**d**) Diffusion map of monocyte gene trajectory, colored by trajectory. (**e**) Expression plots of the T2 and T3 gene signatures between treatments. Expression levels were compared by Wilcoxon rank-sum test with Bonferroni adjustment, whereby **** indicates p<.0001.

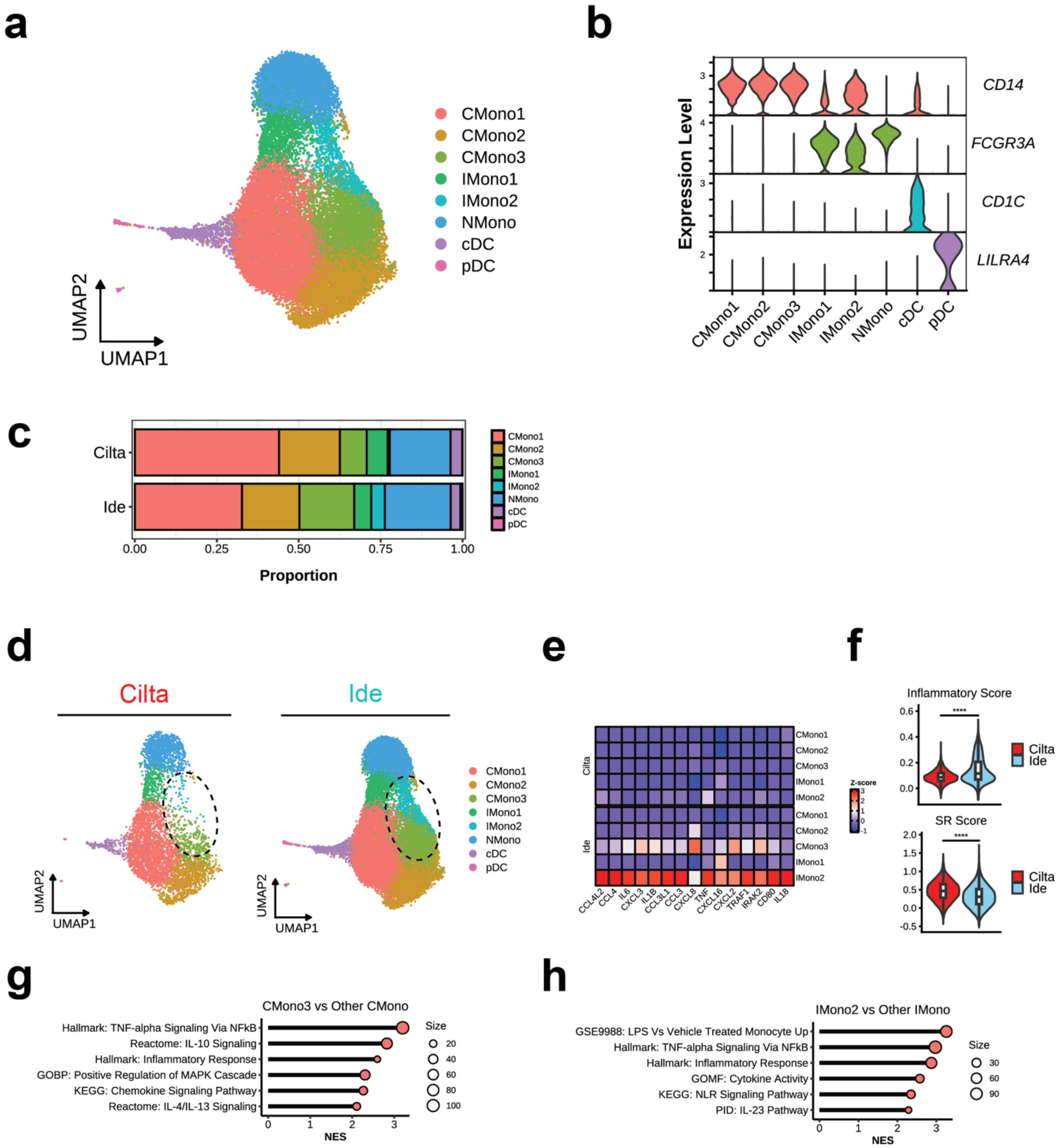
a
CMono1
CMono2
CMono3
IMono1
IMono2
NMono
cDC
pDC
UMAP2
UMAP1
b
Expression Level
CD14
FCGR3A
CD1C
LILRA4
CMono1 CMono2 CMono3 IMono1 IMono2 NMono cDC pDC
c
Cilta
Ide
0.00 0.25 0.50 0.75 1.00
Proportion
d
Cilta
Ide
e
Z-score
CCL4L2 CCL4 IL6 CXCL3 IL1B CCL3L1 CCL3 CXCL8 TNF CXCL16 CXCL2 TRAF1 IRAK2 CD80 IL18
f
Inflammatory Score
SR Score
g
CMono3 vs Other CMono
Hallmark: TNF-alpha Signaling Via NFkB
Reactome: IL-10 Signaling
Hallmark: Inflammatory Response
GOBP: Positive Regulation of MAPK Cascade
KEGG: Chemokine Signaling Pathway
Reactome: IL-4/IL-13 Signaling
Size
NES
h
IMono2 vs Other IMono
GSE9988: LPS Vs Vehicle Treated Monocyte Up
Hallmark: TNF-alpha Signaling Via NFkB
Hallmark: Inflammatory Response
GOMF: Cytokine Activity
KEGG: NLR Signaling Pathway
PID: IL-23 Pathway
Size
NES

**Figure S15: Phenotypic and transcriptomic characteristics of post-infusion myeloid cells in the peripheral blood.** (**a**) UMAP plot of post-infusion endogenous myeloid cells, colored by phenotype. (**b**) Expression plots of key genes used to annotate myeloid phenotypes. (**c**) Bar plot depicting phenotype proportions, split by treatment. (**d**) UMAP plot of post-infusion endogenous monocytes, colored by phenotype. (**e**) Heatmap depicting scaled expression of inflammatory DEGs found to be upregulated by the CMono3 and IMono2 subsets, grouped by subset and split by treatment. (**f**) Violin plots contrasting inflammatory and SR monocyte gene signatures between post-cilta-cel and post-ide-cel monocytes. Expression levels were compared by Wilcoxon rank-sum test with Bonferroni adjustment, whereby **** indicates $p<.0001$. (**g**) Dot plot of pathways enriched on DEGs between the CMono3 subset and other CMono subsets. (**h**) Dot plot of pathways enriched on DEGs between the IMono2 subset and the IMono1 subset. Abbreviations: CMono, conventional monocyte; IMono, intermediate monocyte; NMono, nonconventional monocyte; cDC, conventional dendritic cell; pDC, plasmacytoid dendritic cell.

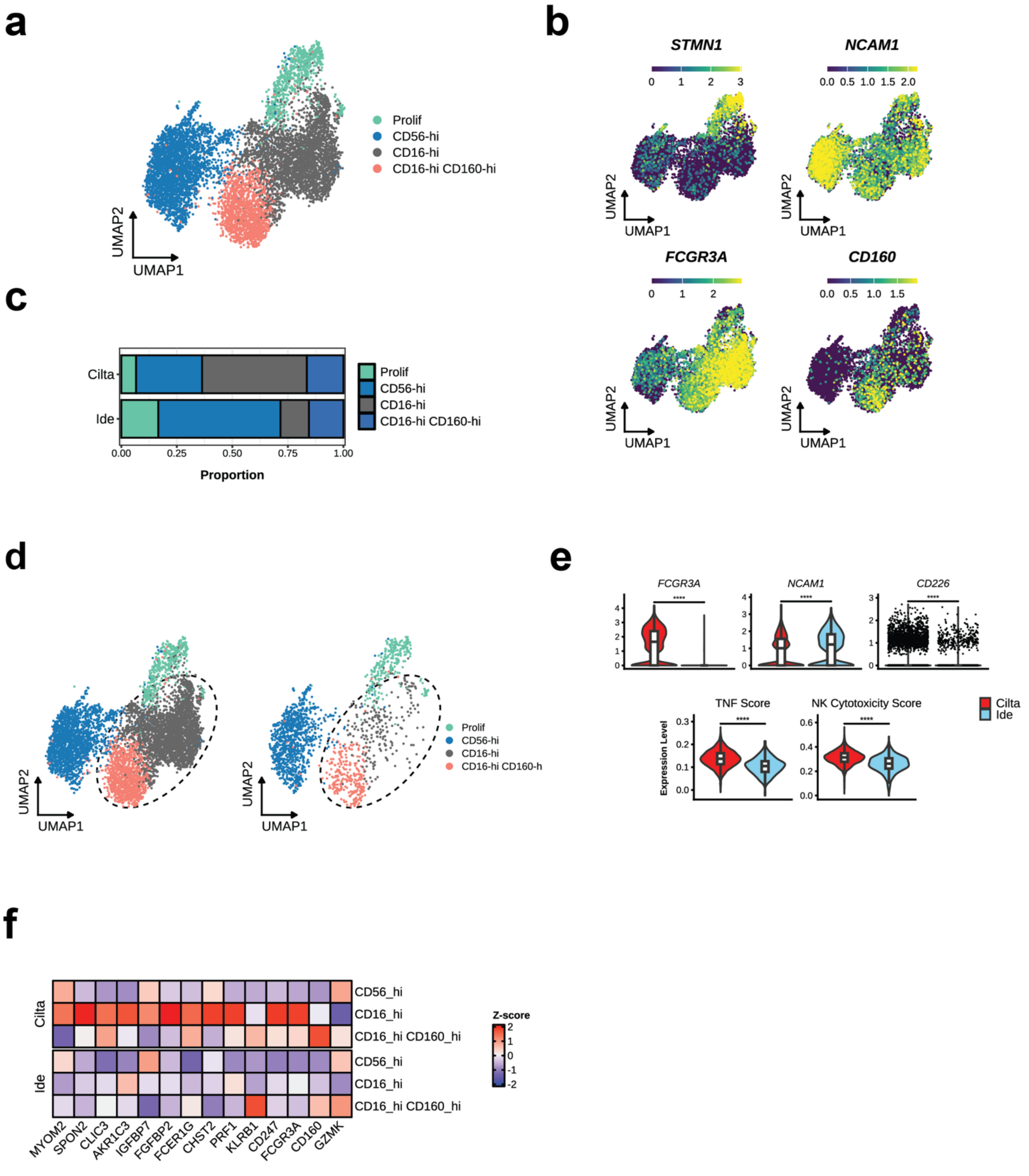
a
b
c
d
e
f
Prolif
CD56-hi
CD16-hi
CD16-hi CD160-hi
UMAP2
UMAP1
STMN1
NCAM1
FCGR3A
CD160
Cilta
Ide
0.00
0.25
0.50
0.75
1.00
Proportion
CD16-hi CD160-h
FCGR3A
NCAM1
CD226
TNF Score
NK Cytotoxicity Score
Expression Level
Z-score
CD56_hi
CD16_hi
CD16_hi CD160_hi
MYOM2
SPON2
CLIC3
AKR1C3
IGFBP7
FGFBP2
FCER1G
CHST2
PRF1
KLRB1
CD247
FCGR3A
CD160
GZMK

**Figure S16: Phenotypic and transcriptomic characteristics of post-infusion NK cells in the peripheral blood.** (**a**) UMAP plot of post-infusion endogenous NK cells, colored by phenotype. (**b**) Expression plots of key genes used to annotate NK phenotypes. (**c**) Bar plot depicting phenotype proportions, split by treatment. (**d**) UMAP plot of post-infusion endogenous NK cells colored by phenotype and split by treatment. (**e**) Violin plots contrasting expression of NK-associated genes (top) and TNF and NK cytotoxicity gene signatures between post-cilta-cel and post-ide-cel NK cells. Expression levels were compared by Wilcoxon rank-sum test with Bonferroni adjustment, whereby **** indicates $p<.0001$. (**f**) Heatmap depicting scaled expression of selected cytotoxic genes, grouped by treatment and split by sample.
